# Simulation-based validation of a method to detect changes in SARS-CoV-2 reinfection risk

**DOI:** 10.1101/2023.09.21.23295891

**Authors:** Belinda Lombard, Harry Moultrie, Juliet R.C. Pulliam, Cari van Schalkwyk

## Abstract

**Background:** Given the high global seroprevalence of SARS-CoV-2, understanding the risk of reinfection becomes increasingly important. Models developed to track trends in reinfection risk should be robust against possible biases arising from imperfect data observation processes.

**Objectives:** We performed simulation-based validation of an existing catalytic model designed to detect changes in the risk of reinfection by SARS-CoV-2.

**Methods:** The catalytic model assumes the risk of reinfection is proportional to observed infections. Validation involved using simulated primary infections, consistent with the number of observed infections in South Africa. We then simulated reinfection datasets that incorporated different processes that may bias inference, including imperfect observation and mortality, to assess the performance of the catalytic model. A Bayesian approach was used to fit the model to simulated data, assuming a negative binomial distribution around the expected number of reinfections, and model projections were compared to the simulated data generated using different magnitudes of change in reinfection risk. We assessed the approach’s ability to accurately detect changes in reinfection risk when included in the simulations, as well as the occurrence of false positives when reinfection risk remained constant.

**Key Findings:** The model parameters converged in most scenarios leading to model outputs aligning with anticipated outcomes. The model successfully detected changes in the risk of reinfection when such a change was introduced to the data. Low observation probabilities (10%) of both primary- and re-infections resulted in low numbers of observed cases from the simulated data and poor convergence.

**Limitations:** The model’s performance was assessed on simulated data representative of the South African SARS-CoV-2 epidemic, reflecting its timing of waves and outbreak magnitude. Model performance under similar scenarios may be different in settings with smaller epidemics (and therefore smaller numbers of reinfections).

**Conclusions:** Ensuring model parameter convergence is essential to avoid false-positive detection of shifts in reinfection risk. While the model is robust in most scenarios of imperfect observation and mortality, further simulation-based validation for regions experiencing smaller outbreaks is recommended. Caution must be exercised in directly extrapolating results across different epidemiological contexts without additional validation efforts.

## Introduction

The COVID-19 pandemic has had catastrophic health, economic, and social impact, directly affecting billions of lives. As of July 2023, the pandemic had resulted in at least 6.9 million deaths globally (1). Five major waves of infections were observed in South Africa. The first wave, driven by the original strain, peaked in mid-2020 and was followed by a second wave driven by the Beta variant towards the end of 2020. The Delta variant drove the third wave, in mid-2021, and the fourth and fifth waves were driven by the BA.1/ BA.2 and BA.4/ BA.5 Omicron sub-variants, at the end of 2021 and May 2022 (2). These waves, coupled with vaccination efforts, have resulted in high levels of seroprevalence and relatively low numbers of reported infections since mid-2022 (3).

Reinfection with SARS-CoV-2 has emerged as a concern, due to waning immunity following infection and imperfect immunity, whereby prior infection does not provide full protection against reinfection (4). Viral evolution also leads to the emergence of new variants, which may increase risk of reinfection.

Understanding the risk of reinfection by SARS-CoV-2 and potential future epidemics with other pathogens which does not result in lifelong immunity, has significance for both individual and public health. At the individual level, awareness of a high risk of reinfection might encourage individuals to take necessary precautions. In the public health context, understanding the risk of reinfection can help health officials make more informed decisions, potentially recommending increased practice of protective measures like hand sanitising and mask-wearing in public spaces, particularly if the reinfection risk is high.

### Modelling studies of SARS-CoV-2 reinfection

Multiple studies, including several modelling studies, have been conducted to assess SARS-CoV-2 reinfection patterns. A Susceptible-Exposed-Asymptomatic-Infectious-Recovered (SEAIR) epidemic model that includes reinfections has been developed and applied to SARS-CoV-2 data in Pakistan, highlighting the importance of understanding reinfections in controlling disease spread (5). A Brazilian study utilised a more complex compartmental disease model, incorporating hospitalisation and deaths to assess the force of reinfections by the P.1 variant confirming that the P.1 variant significantly contributed to a surge of reinfections (6).

In addition to these modelling studies, a catalytic model was developed to monitor SARS-CoV-2 reinfection trends in South Africa and identified population-level changes in the risk of reinfection (7). The catalytic model estimates the number of expected reinfections through time under the assumption of a constant reinfection risk. Using Monte-Carlo Markov Chains (MCMC), the model was fitted to data on observed reinfections during the first two waves of SARS-CoV-2 in South Africa. The estimated reinfection hazard coefficient was then used to project reinfection numbers during the two subsequent waves. Notably, the number of observed reinfections remained within the projection interval during the third (Delta) wave but diverged early in the fourth (Omicron BA.1/BA.2) wave, providing the first evidence of the variant’s ability to evade immunity from prior infections (7).

### Imperfect representation of the real-world in models

Observed infection data is not always fully representative of the real world (8) due to factors such as undetected asymptomatic or mild cases, inaccessible testing centres, and variable testing behaviour (9,10). Testing behaviour within a population may also vary in response to the number of reported infections (10).

These factors, coupled with underreporting of SARS-CoV-2 infections and COVID-19 mortality, can skew the assumed number of people at risk of reinfection (11). Therefore, methods for assessing reinfection risk need to be evaluated to determine how these biases impact performance (12). Inaccurate models could lead to incorrect predictions and conclusions regarding changes in reinfection risk, which may have negative effects on public health and the economy (12,13).

Simulation-based validation can be used to validate a model’s robustness to specific assumptions that are likely to be violated in the real world. To do this, simulated datasets are generated from a variety of processes that violate the assumptions to be tested. The model’s performance is then evaluated based on these data, which emulates a more real-world situation. Performance is assessed across a wide range of assumptions and parameter values. When these analyses reveal that certain parameters or assumptions affect the model’s performance, it is crucial to refine the modelling approach to account for these factors (14).

In this study, we applied simulation-based validation and associated sensitivity analyses to evaluate the performance of the catalytic model presented in (7), by introducing different biases that represent real-world phenomena to simulated data and assessing whether the method can be used to correctly evaluate reinfection trends under these conditions.

## Materials and Methods

### The catalytic model

The catalytic model was developed to assess changes in reinfection risk by SARS-CoV-2 accounting for both the changing underlying number of individuals who have already been infected and the changing infection risk through time (7). In this model, a reinfection is defined as an individual having two positive tests for SARS-CoV-2 at least 90 days apart. This delay is introduced to align with the referenced paper, to ensure that successive positive tests result from reinfection rather than prolonged viral shedding. Consequently, the model sets the risk of reinfection at zero for the first 90 days, and thereafter, it is proportional to the 7-day moving average of observed infections.

The probability of a positive test for SARS-CoV-2 by day *x* after *t* is given by the equation:

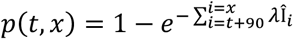

where *λ* is the reinfection hazard coefficient and Î_*i*_ is the 7-day moving average of the total number of infections (both primary infections and reinfections) on day *i*.

The expected number of cases where the first positive test was on day *t* with a detected reinfection by day *x* is given by *I*_*t*_*p*(*t, x*), where *I*_*t*_ is the number of putative primary infections reported on day *t*.

Thus, the expected number of reinfections by day *x* can be expressed as:

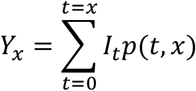

### Model fitting and projection

The catalytic null model, which assumes a constant reinfection hazard coefficient, can be fitted to the number of observed reinfections up until a defined ‘fitting date*’*. The time before this fitting date is referred to as the ‘fitting period*’*.

In this process, two parameters were fitted using Metropolis-Hastings Monte Carlo Markov Chains (MCMC): the reinfection hazard coefficient (*λ*) and the negative binomial dispersion parameter (*κ*), assuming that the number of reinfections follows a negative binomial distribution. In the simulation-based validation conducted in this study, four Markov chains were run (each with a random starting value) with 10 000 iterations in each chain. The first 4000 iterations of each chain were discarded as burn-in.

A joint posterior distribution of 1600 parameter sets were obtained from the Markov chains that we obtained in the fitting procedure, by selecting every 15^th^ sample from the joint distribution. Each sample in the joint posterior distribution was used to simulate 100 stochastic realisations of expected daily reinfections. The stochastic realisations were used to obtain a 95% uncertainty interval for the fitting period, and a 95% credible interval for the ‘projection period’ (the time after the fitting date) under the null model.

We used simulation-based validation to evaluate the robustness of using a metric of five consecutive data points that lie above the projection interval as an indicator of an increase in reinfection risk. We also assessed the effect of violating certain assumptions on the model’s convergence during the fitting period, and on the proportion of points that lie above the projection interval after a change in reinfection risk.

### Simulation-based validation

The simulation-based validation started with the construction of a simulated dataset of primary infections representing a world in which all SARS-CoV-2 infections are observed, and no deaths occur, therefore every infected individual becomes eligible for reinfection after a 90-day period.

Figure 1 shows the simulated dataset containing the number of primary infections per day. This dataset of primary infections is based on the number of observed infections in South Africa through July 2021. Specifically, we generated a simulated time series of primary infections by taking the seven-day moving average of first infections from the previously published South African data (15). We increased the seven-day moving average by a factor of 5, then took a negative binomial sample around this mean with a shape parameter of 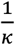, where *κ* ≈ 0.27 was the median of the posterior sample from (7).For dates at the beginning of the time series for which a seven-day moving average could not be calculated, the observed count was inflated by a factor of five and used as the mean for the negative binomial draw.

**Figure 1.**
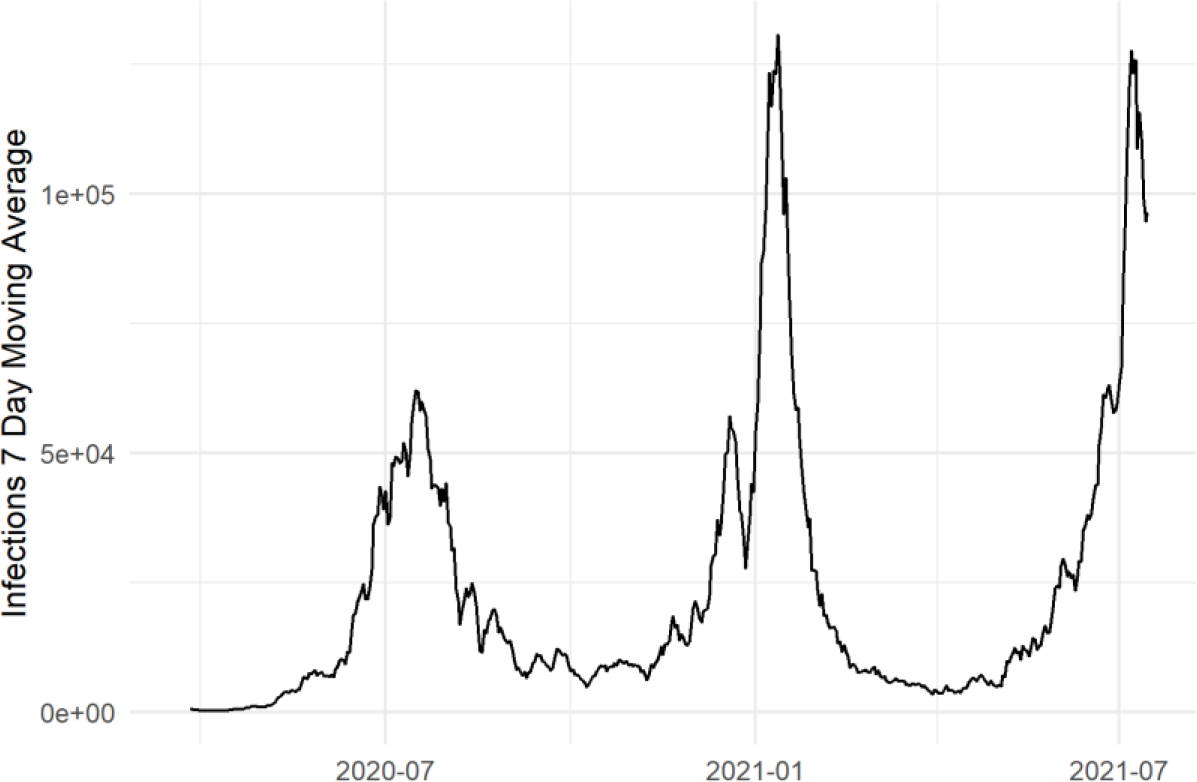
Simulated underlying daily infections that represents three waves of COVID-19 in South Africa. The simulated data is based on the observed cases in South Africa up to 15 June 2021 by scaling the number of cases and adding noise to the number of infections.

### The simulated scenarios

We utilised five scenarios to assess the model’s robustness and reliability of our chosen metric for detection of a change in reinfection risk. The first four scenarios depict an increasingly more realistic model world, with the fifth scenario considering a more complex description of varying observation probability as described in Table 1.

**Table 1.**
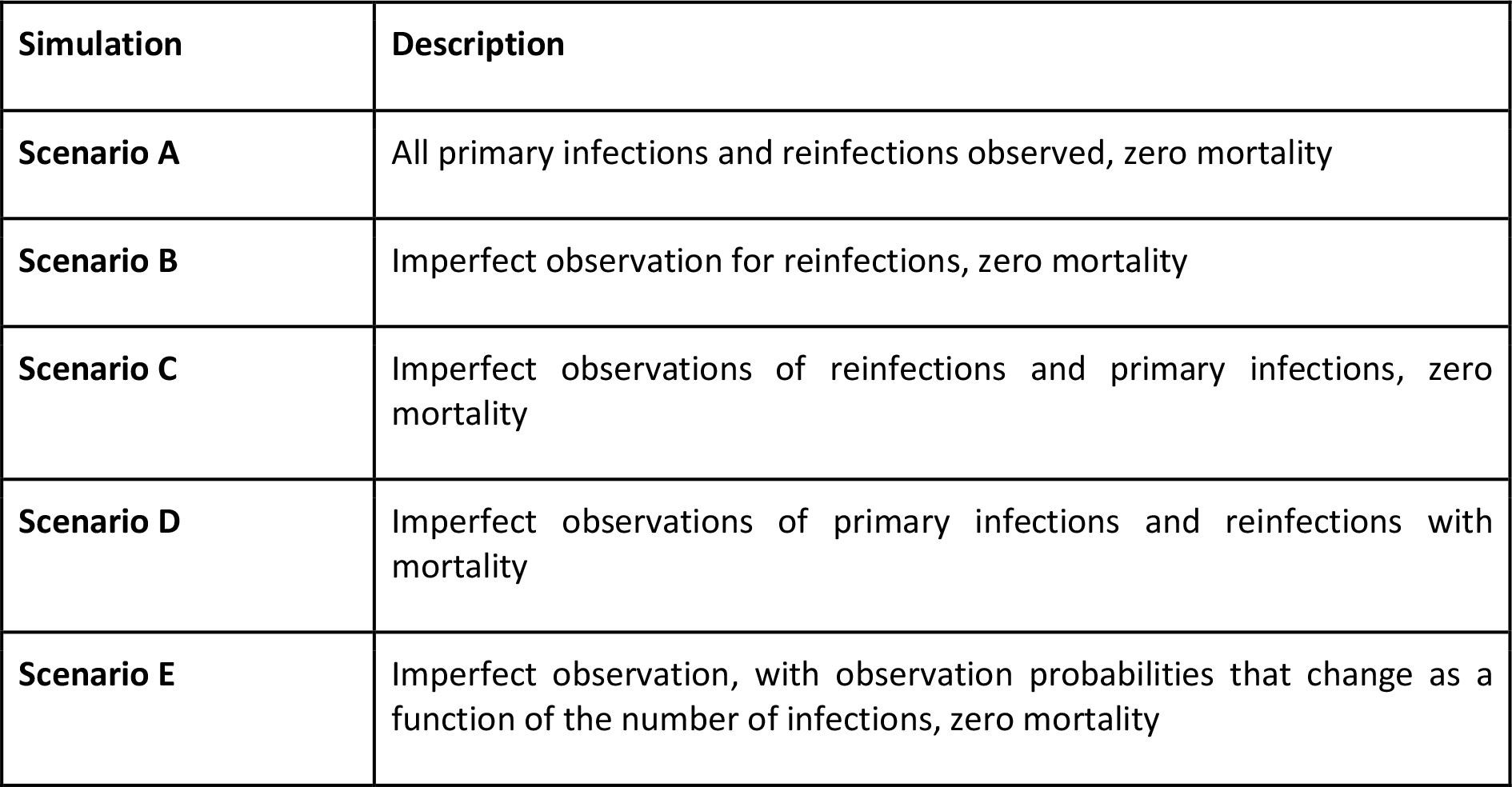
Summary of each scenario used in the simulation-based validation, illustrating a progression towards more realistic data generation processes from Scenario A to D and Scenario E covering a scenario of varying observation probability without mortality.

| Simulation | Description |
| --- | --- |
| **Scenario A** | All primary infections and reinfections observed, zero mortality |
| **Scenario B** | Imperfect observation for reinfections, zero mortality |
| **Scenario C** | Imperfect observations of reinfections and primary infections, zero mortality |
| **Scenario D** | Imperfect observations of primary infections and reinfections with mortality |
| **Scenario E** | Imperfect observation, with observation probabilities that change as a function of the number of infections, zero mortality |

**Table 2.** Parameters varied in Scenario E with corresponding values.

| Parameter | Values |
| --- | --- |
| Scale ($\sigma$) | {1; 1.2; 1.5} |
| Maximum observation probability for primary infections ($P_1^{max}$) | {0.2; 0.3; 0.4; 0.5} |
| Minimum observation probability for primary infections ($P_1^{min}$) | {0.1; 0.2; 0.3; 0.4; 0.5} such that<br>$P_1^{min} < P_1^{max}$ |
| Maximum observation probability for reinfections ($P_2^{max}$) | {0.2; 0.3; 0.4; 0.5} such that<br>$P_2^{max} \geq P_1^{max}$ |
| Minimum observation probability for reinfections ($P_2^{min}$) | {0.1; 0.2; 0.3; 0.4; 0.5} such that<br>$P_2^{min} < P_2^{max}$ and $P_2^{min} \geq P_1^{min}$ |
| Steepness ($s$) | {0.00005; 0.0001} |
| Midpoint ($x_m$) | {30000; 40000; 50000} |

As part of the simulation-based validation, we sought to determine when a change in the hazard coefficient may be detected in the model, given the real-world evidence that certain variants carry higher risks of reinfection (7). When applying the model to the actual reinfection data in South Africa, the reinfection hazard coefficient during the Omicron wave was estimated to be higher than in the previous waves (7). Given this, we generated ‘true’ datasets for each scenario where the reinfection hazard coefficient either remains the same or is higher after a specific date. These datasets allow us to test the catalytic model against both the situation where there is a wave driven by a variant with a higher risk of reinfection, and the situation where the reinfection risk remains constant. This assessment provides insights into the model’s sensitivity and specificity of detecting a change in reinfection risk under different scenarios.

For data generation, a scale parameter was introduced after a ‘scale date’ (which we used as 1 May 2021) that is used to represent an Omicron-like wave, which varied from 1 (representing no change in reinfection risk) to 3 by steps of 0.1:

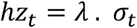

where *hz*_*t*_ is the modified hazard coefficient used to calculate reinfections on day t, *λ* is the reinfection hazard coefficient (obtained from the median of the posterior distribution of the fitted reinfection hazard coefficient in (7)) and *σ*_*t*_ is a modifier on the hazard defined as

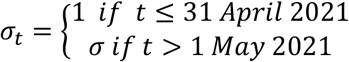

We used *σ* to represent an increase in reinfection risk to represent the Omicron wave and varied *σ* ≥ 1 in the different scenarios.

Detailed simulations for each scenario, including calculations and parameter adjustments, are described below.

### Scenario A: Perfect observation and no mortality

The rationale for this approach is to establish a baseline, where we assess how our model performs when every infection of SARS-CoV-2 is observed, and no mortality occurs. This will determine the model’s ability to converge when all cases are observed and its ability to detect changes in the risk of reinfection with different magnitudes of these changes. In the simulated data, infected individuals become eligible for reinfection 90 days after their first infection.

The simulated primary infection dataset described above is used to calculate reinfections as follows:

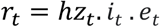

where *hz*_*t*_ is the modified reinfection hazard coefficient, *i*_*t*_ is the number of underlying primary infections on day *t* and *e*_*t*_ represents the number of people that are eligible for reinfection on day *t* and is calculated as follows:

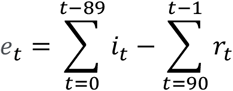

The number of people eligible for reinfection, *e*_*t*_, is calculated by subtracting the number of people who have already been reinfected with SARS-CoV-2 from those who had a primary infection at least 90 days ago, by that day.

The only parameter that is varied in this scenario is the scaling of the reinfection hazard (*σ*), introduced at the start of the third wave. We varied this scale parameter from 1 to 3 in steps of 0.1 to determine the value of the scale parameter at which the observed data points no longer fit within the projection interval produced by the model.

The data simulated for this scenario can be seen in Figure S1.

### Scenario B: Imperfect observation of reinfections

Scenario B introduces the first potential factor that may bias the detection of a change in reinfection hazard coefficient, namely the challenge where not every reinfection is observed, which mirrors actual epidemiological settings where not every case is reported or observed. By introducing this variable, we aim to evaluate the impact of imperfect observation of reinfections on the model’s robustness.

The number of underlying reinfections is calculated as in Scenario A as follows:

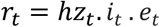

We then add an observation probability to calculate the number of observed reinfections, which is represented as:

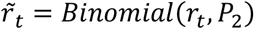

where *P*_2_ is the varying observation probability and 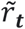 is the number of observed reinfections. We use the binomial distribution to calculate the number of observed cases for added noise which is present in a real-world scenario. The observed number of people eligible for reinfection is represented by 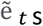 and is based on the number of observed reinfections, instead of the underlying reinfections, and is calculated as:

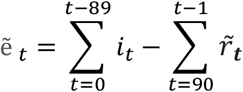

The underlying number of people eligible for reinfection, *e*_*t*_, is calculated as in Scenario A and is used as the basis for determining the underlying number of reinfections, as shown above; however, when the catalytic model is applied to the simulated data, the observed number of eligible infections is used for fitting.

Alongside the scale parameter (*σ*) like in Scenario A, we also vary the reinfection observation probability, *P*_2_, from 0.1 to 0.5 in increments of 0.1.

The data simulated for this scenario can be seen in Figure S2.

### Scenario C: Imperfect observation of reinfections and primary infections

In real-world settings, both primary infections and reinfections can go unnoticed, under-reported, or unrecorded. Thus, it is necessary for any model to handle the imperfect observation of both primary infections and reinfections, each with a specified observation probability. Scenario C introduces this added complexity, to determine the model’s performance when confronted with imperfect observation of primary infection and reinfections.

Before calculating the number of reinfections, we must first adjust for the imperfect observation of primary infections:

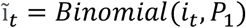

Here, *P*_1_ denotes the observation probability for primary infections, which is varied in this scenario from 0.1 to 0.5 in increments of 0.1. We again utilised a binomial distribution draw to add noise to the number of reinfections consistent with the approach for primary infections. Because the dataset for analysis in the real world would be based only on first observed infections, detected reinfections would only be possible in the individual’s whose first infection was detected; we therefore use the observed primary infections in calculation of the simulated time series for reinfections.

Once the number of observed primary infections is determined, the number of underlying reinfections (within this open cohort) per day is calculated as follows:

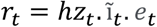

where ĩ_*t*_ is number of the observed reinfections on day t, and *e*_*t*_ is calculated as:

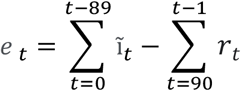

Like Scenario B, an observation probability is added to the number of reinfections using the binomial distribution:

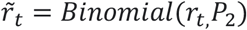

Here, the observed number of people eligible for reinfection is determined as follows:

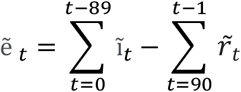

Three parameters are varied in this scenario: primary infections observation probability (*P*_1_), reinfections observation probability (*P*_2_) and the scale parameter (*σ*).

The data simulated for this scenario can be seen in Figure S3.

### Scenario D: Imperfect observations of primary infections and reinfections with mortality

Mortality from individuals with a primary infection significantly impacts the number of people eligible for reinfection, and if not factored into a model, it could result in overestimation. In the fourth scenario, we factor in those who have died from a primary infection, which influences the cohort of individuals susceptible to reinfection, giving a more refined perspective on SARS-CoV-2 transmission dynamics. The number of deaths resulting from observed primary infections is calculated as:

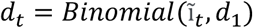

In this equation, *d*_1_ is the probability of dying, a parameter that is being varied in this scenario. The number of observed primary infections, ĩ_*t*_ is, is calculated in the same way as in Scenario C.

The number of underlying reinfections per day is calculated as in Scenario C:

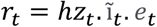

As in Scenario C, an observation probability is introduced to the number of underlying reinfections to calculate the number of observed reinfections, 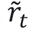.

In contrast to previous scenarios, however, the number of people deemed eligible for reinfection is adjusted by factoring in those who died from a primary infection and thus cannot be reinfected. This is calculated as follows:

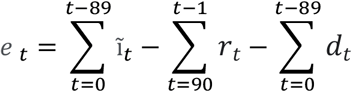

The number of observed people who remain susceptible for reinfection is then calculated as:

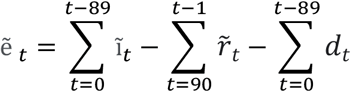

The parameters that are being varied in this scenario are *d*_1_, the probability of dying of the primary infection (*d*_1_ = 0.001, 0.01, 0.05), the scale for the last wave’s hazard coefficient *σ* (as with Scenario A), the observation probability for reinfections *P*_2_ (as per Scenario B) and the observation probability for primary infections *P*_1_ (consistent with Scenario C).

### Scenario E: Imperfect observation, with observation probabilities that change as a function of the number of infections

The rationale for this scenario is to reflect a real-world setting, where potential changes in testing behaviour could be influenced by the perceived infection prevalence and/or the saturation of testing services during a surge. Our model would then be more adaptive and aligned with a more dynamic real-world scenario.

In Scenario E, we introduce dynamic observation probabilities (*P*_1_, and *P*_2_,) as a function of the number of underlying infections. At times where the incidence is high, the observation probabilities might be lower since individuals are less likely to test for infection or test availability may be limited. Thus, we incorporate this into the model by varying the observation probability with the inverse of the logistic function, making it a function of the number of underlying infections on day t. We calculate the observation probabilities on day t, *f*(ĩ_*t*_), as:

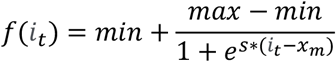

In this equation, *min* and *max* are represented by the minimum and maximum observation probabilities respectively, *s* is the steepness parameter and *x*_*m*_ is the ‘mid-point’. Figure 2 visually depicts *f*.

**Figure 2.**
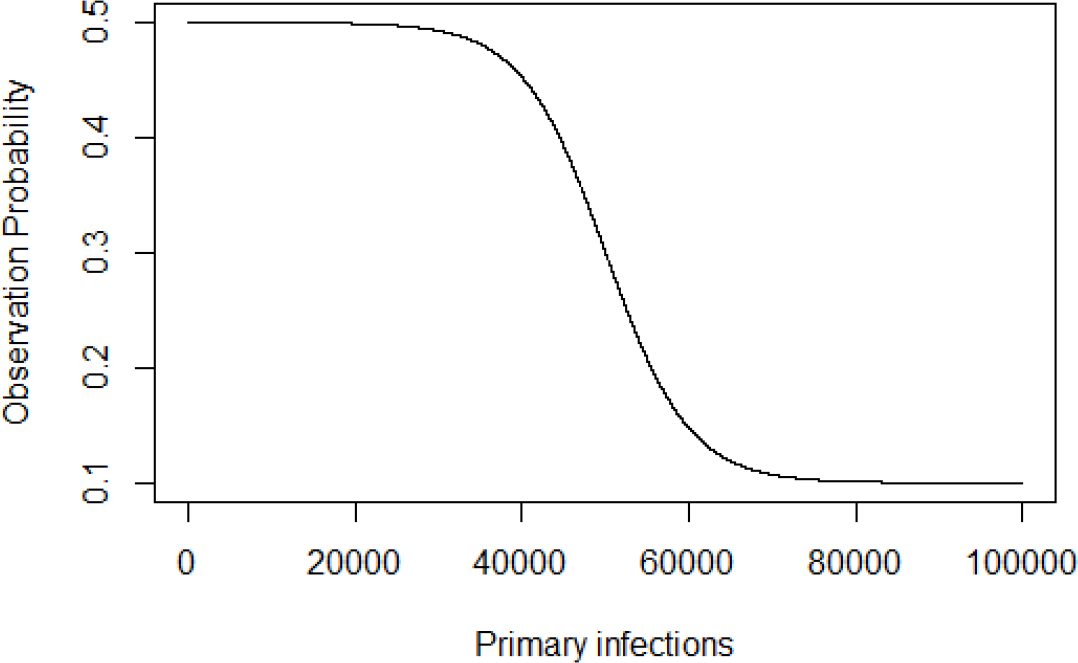
The function (a variation of the logistic function) plotted which illustrates changes in the observation probability with shifts in the number of underlying primary infections on that day.

Using this function, the observation probabilities, and then the number of observed primary infections and reinfections are calculated. The only variables that are being changed between primary infections and reinfections are the minimum and maximum observation probabilities, since behaviour depends on the number of underlying primary infections and not the number of underlying reinfections (in the case of reinfections).

The maximum and minimum observation probabilities for reinfections (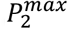 and 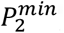) such that

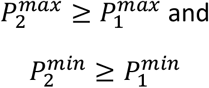

where 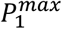 and 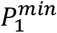 are the observation probabilities for primary infections, respectively.

We hypothesise that people who are reinfected are more likely to get tested for SARS-CoV-2 since they tested for their primary infections.

We excluded parameter sets where 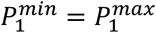 and 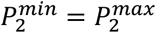 as this would replicate Scenario C.

Multiple parameters are varied in this scenario: the maximum and minimum observation probability for primary infections (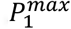 and 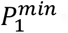), the maximum and minimum observation probability for reinfections (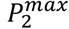 and 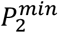), the steepness of the function (*s*), the midpoint of the function (*x*_*m*_), and the scale (*σ*).

In Imperfect observation, with observation probabilities that change as a function of the number of infections, each of the parameters that are being varied in Scenario E are described and summarised with its corresponding values that are used in the simulation-based validation process.

### Evaluating model performance

We assessed performance of the catalytic method for detecting increases in the reinfection risk across Scenarios A through E. For each scenario, data were simulated specific to the parameter set that characterises the scenario. We applied the fitting procedure and projection to each simulated dataset as described above to estimate a 95% projection interval for reinfections per day in each simulated dataset.

The fitting and projection process was repeated 20 times for each scenario, each time using different seed values. These varied seeds generated different sets of random numbers which were used in three key processes:

1. The data generation process where binomial processes were used to generate the number of observed cases and mortality.
2. The MCMC fitting procedure when calculating the proposer value, which is drawn from a normal distribution and accepted or rejected by the MCMC sampler within the Markov chain.
3. The model projection process when calculating the stochastic realisations.

We assessed parameter convergence and model fit during the fitting period, then applied a set of metrics to measure the impact of different scenario definitions on the model performance. These metrics included: assessing the first cluster of reinfections above the projection interval during the projection period, determining the proportion of infections above the projection interval during the projection period, and evaluating the specificity of detecting simulated changes in reinfection risk.

The model was implemented, and the simulation-based validation was conducted in the R Statistical Programming Language [version 4.3.1 (2023-06-16)] (16). The code is available on Github at https://github.com/SACEMA/reinfectionsBelinda.

#### Parameter convergence

In each scenario, the MCMC sampler of the catalytic model was used to fit the model parameters. These parameters include *λ*, which is the reinfection hazard coefficient, and *κ*, the binomial dispersion parameter. After the fitting procedure, the convergence of *λ* and *κ* was measured using Gelman-Rubin convergence diagnostics, which relies on a potential scale reduction factor (PSFR) commonly used to measure convergence in Markov chains (17). The Gelman-Rubin convergence diagnostic is a ratio that compares the between-chain and within-chain variances. When the difference between the respective variances is large, it indicates non-convergence (17,18). A value below 1.1 indicates that the parameter converged, and a value above 1.2 indicates non-convergence (17). To determine parameter convergence, we measured the proportion of runs where convergence was achieved (i.e., where PSRF ≤ 1.1 for both *λ* and *κ*) when considering different scenario definitions involving observation probabilities and mortality.

#### Exclusion of non-converging runs or poor model fit

When *κ* does not converge, the projection bands are narrow which could lead to more observed reinfections falling outside the projection interval. Such an outcome could lead to incorrect conclusions regarding trends in the risk of reinfection. Similarly, it is crucial for *λ* to converge to ensure reliable predictions. We therefore excluded runs where either *λ* or *κ* did not converge from our analysis.

Additionally, for each run of the scenario definitions, we tracked the presence of clusters of data points that fell outside the 95% projection interval for the 7-day moving average of reinfections during the fitting period. Where such a cluster existed, it implied that the model inaccurately represents the patterns seen in the simulated data. In addition to excluding non-converging runs, we also excluded runs for which a cluster of either five consecutive observed values for the 7-day moving average of reinfections fell above the projection interval or ten consecutive values fell below the 95% 7-day moving average projection interval during the fitting period (before the fitting date, 28 February 2021).

#### First cluster of reinfections above the projection interval

The first cluster of five consecutive points above the projection interval (*D*_*first*_) is a metric that can easily be used for real-time detection of a change in the hazard coefficient. Therefore, it is important to test how well this metric performs under different scenarios.

For runs that were not excluded, we tracked the timing of the first occurrence of five data points (of the 7-day average of observed reinfections) above the 95% 7-day moving average projection interval during the projection period after introducing *σ* (i.e., after 1 May 2021), designated as *D*_*first*_ (where *D*_*first*_ is the fifth day of such a cluster). The presence of *D*_*first*_ indicates a possible increase in reinfection risk. In our simulated data, where we have introduced an increasing hazard coefficient (*σ* > 1), we used *D*_*first*_ to assess the magnitude of change required for our approach to detect it. Additionally, we evaluated how quickly the approach identifies the increase in reinfection risk for different scenarios, identifying any delays from introduction of the increase to its detection (*D*_*first*_). As a summary metric, we calculated the median of *D*_*first*_ after excluding non-converging runs and runs with clusters of reinfections outside the projection interval in the fitting period, as described above.

#### Proportion of infections above the projection interval

The impact of the different processes, represented by different scenarios, on the model’s performance was also measured by determining the proportion of days when the 7-day moving average of observed reinfections fell above the 95% 7-day moving average projection interval after the introduction of *σ*. In cases where there was no increase in the risk of reinfection, we expected that the number of “observed” (simulated) reinfections exceeding the 95% projection interval would be less than 2.5% of the days in the projection period. If more than 2.5% of the daily observed reinfections values fall above the projection interval, it indicated either a successfully detected (when *σ* > 1) or a false positive detection (when *σ* = 1). As a summary metric, we calculated the median of this proportion across runs that were not excluded by the criterion described above.

Measuring the proportion of infections above the projection interval helped us in assessing the magnitude of change in reinfection risk likely to be detected by the method and the effect of the potential biases we examined. It is important to note that this measurement does not enable us to assess real-time performance of the method but is a general indicator of robustness.

#### Specificity

Specificity is a proportion used to measure the approach’s reliability when there is no change in the risk of reinfection. We measured the specificity for each scenario definition where *σ* = 1 by calculating it across the 20 runs as

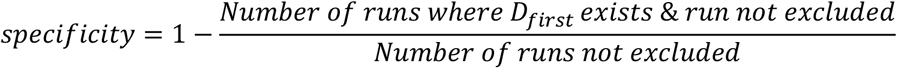

The runs not excluded are those that converged and did not have a large cluster of observed reinfections outside the projection interval during the fitting period, as described above. High specificity indicates that false positive detections of a change in reinfection risk are unlikely.

## Results

### Parameter convergence

In Scenarios A to D, observation probabilities (*P*_1_ and *P*_2_) were varied but kept constant throughout each run (Figure S4, Figure S5). After running the data generation process, the MCMC fitting procedure and the model projection with 20 different seeds, the convergence diagnostics were below 1.1 for *λ* (the reinfection hazard coefficient) and *κ* (the negative binomial dispersion parameter) across most scenario definitions. However, when *P*_1_ and *P*_2_ were lower (for example, at *P*_1_ = 0.1 and *P*_2_ = 0.2), the proportion of runs that converged ranged between 0.65 and 0.85. At the lower extremes for *P*_1_ and *P*_2_ (*P*_1_ = 0.1 and *P*_2_ = 0.1), the proportion of runs that converged ranged between 0.05 and 0.3, with an increase observed as the probability of dying from a primary infection (*d*_*t*_) decreased (Figure S4, Figure S5).

In Scenario E, the observation probabilities for primary and reinfections varied depending on the number of underlying primary infections for a given day. When 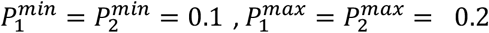, and the steepness of the function (how quickly the observation probability declines with an increase of underlying primary infections) *s* was low, along with a low midpoint (middle of the decline in observation probabilities) *x*_*m*_, most of the runs did not converge (Figure S6, Figure S7). These scenarios corresponded to relatively few observed primary infections and consequently, few generated reinfections. In other cases, i.e., at higher values of 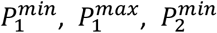 and 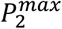, more than 75% of runs converged (Figure S8 and Figure S9).

### Exclusion of non-converging runs or poor model fit

In Scenario A, all runs were included in the analysis. In Scenario B, two runs were excluded due to a cluster of five consecutive points falling above the projection interval during the fitting period. These exclusions occurred in cases where *P*_2_ = 0.1 and *P*_2_ = 0.4, out of a total of twenty runs. In Scenarios C and D, when *P*_1_ = 0.1 and *P*_2_ = 0.1, the majority of runs were excluded due to non-convergence and/or a cluster of five consecutive points above the projection interval during the fitting period. The exceptions were with *d*_1_ = 0.01 where one out of the 20 runs were included. When *P*_1_ = 0.1 and *P*_2_ = 0.2, between three and seven of 20 runs were excluded for each scenario definition. For the rest of the values of *P*_1_ and *P*_2_ (when *P*_1_ > 0.1 and *P*_2_ > 0.2), at most two of 20 was excluded due to non-convergence.

In Scenario D, there were 1500 runs (resulting from the combination of 20 stochastic repeats x three *d*_1_ parameters x 25 combinations of *P*_1_ and *P*_2_) where *σ* = 1, and from these runs, 81 were excluded due to non-convergence, and an additional 107 runs were excluded because of a cluster of five consecutive points falling above the projection interval during the fitting period. No clusters of ten consecutive points below the projection interval during the fitting period were observed in Scenario D.

In Scenario E, there were 6000 runs (20 stochastic repeats x two midpoints parameters x three steepness parameters x 50 combinations of 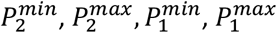) where *σ* = 1. Only 83 of these runs were excluded because of non-convergence, with an additional 1386 being excluded due to clusters of five consecutive points above the projection interval during the fitting period. Additionally, 1701 runs were excluded due to clusters of ten consecutive points falling below the projection interval during the fitting period. One such scenario definition can be seen in Figure S10, where a cluster of 10 consecutive observed reinfections was below the projection interval during the fitting period. In these excluded runs, the simulated data did not match observed trends in South African data, where peaks in primary infections and reinfections were temporally correlated.

### First cluster of reinfections above the projection interval

The first cluster of reinfections above the projection interval, Figure 3 *D*_*first*_, is illustrated in Figure 3 showing the median timing across the five scenarios based on their respective definitions.

**Figure 3.**
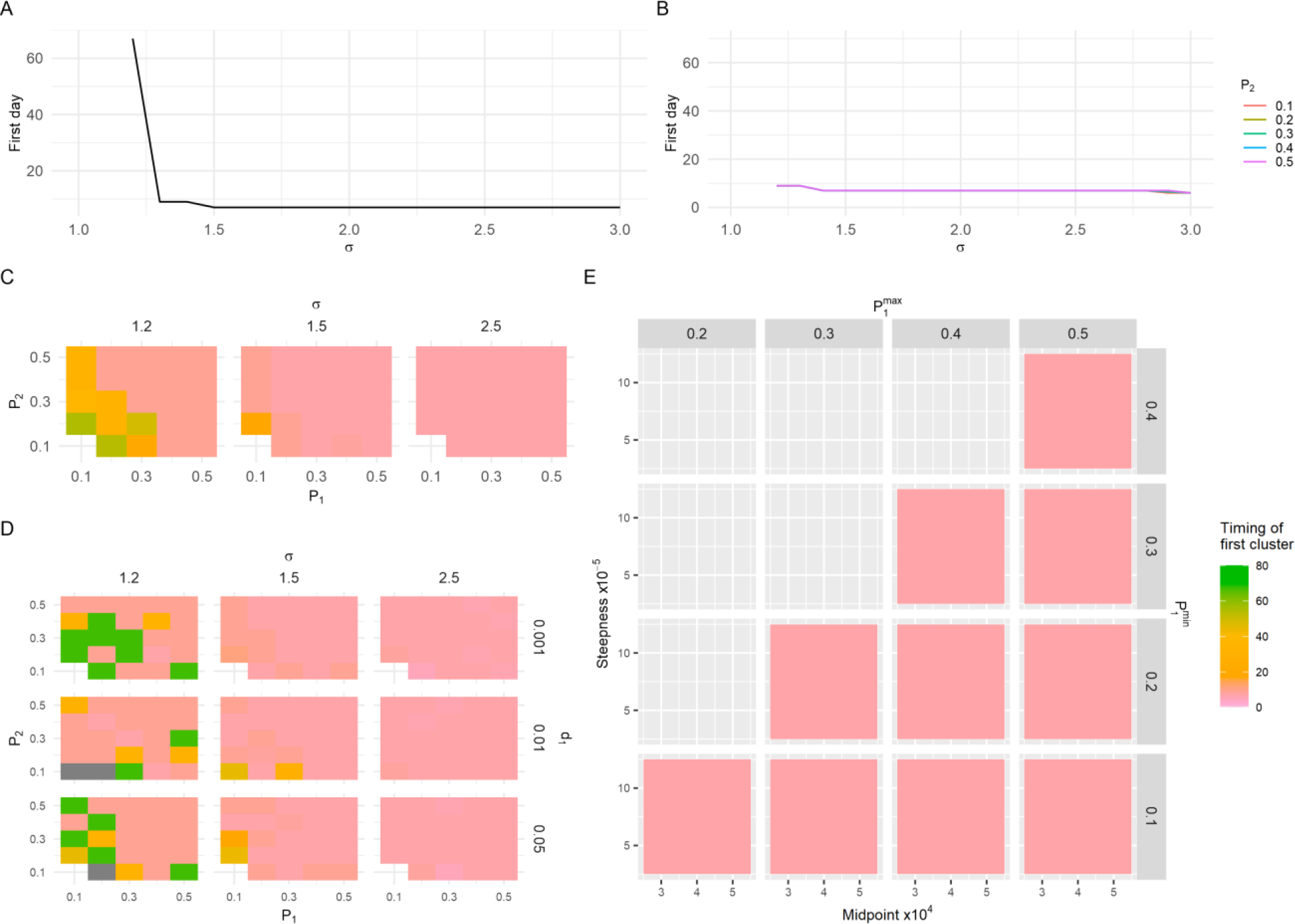
Timing of the first five consecutive points outside the projection interval after an increase in reinfection risk has occurred. Panels A-E represents Scenario A-E respectively. In A, all cases are observed, and no mortality occurs; B represents imperfect observation of reinfections; C represents imperfect observation of primary infections and reinfections; D represents imperfect observation of primary infections and reinfections with added mortality; and E represents observation probabilities that vary with the number of underlying infections without mortality. In E, σ=1.5 and 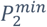 and 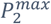 are 0.4 and 0.5 respectively. Plots for other Scenario E definitions are shown in Figure S11. The gaps in C and D represent scenario definitions where results of all runs were excluded because of non-convergence or a cluster outside the projection interval during the fitting period.

In Scenarios A through D, for most of the scenario definitions, the median of *D*_*first*_ = 7 when *σ* ≥ 1.5, indicating the change in reinfection risk is detected very soon after it is introduced in the data. In Scenarios C and D, when *σ* = 1.2, the median of *D*_*first*_ over the 20 runs was slightly higher for most scenario definitions, but the change in risk of reinfection was still detected (i.e., *D*_*first*_ exists for all non-excluded runs). Generally, lower values of *P*_1_ and *P*_2_ were associated with slightly higher median values of *D*_*first*_. In Scenario D, when the probability of mortality after experiencing an observed primary infection, *d*_1_, is lower, the median *D*_*first*_ was also slightly higher.

In Scenario E, when *σ* > 1.2, the change in reinfection risk was detected for all non-excluded runs. However, when the number of observed cases (primary infections and reinfections) was lower, the median *D*_*first*_ was higher (extending to around 40 days) (as shown in Figure S11). At *σ* = 1.2, *D*_*first*_ was absent for eight out of 2833 runs, all at low observation probabilities of primary infections and reinfections.

### Proportion of infections above projection interval

In all the scenarios (A to E), the proportion of data points above the projection interval gradually increased as the scale parameter (proportional increase in risk of reinfection), *σ* increased up to a point after which the proportion stabilised around 0.973 (Figure 4 A-E). The value 0.973 was obtained when 73 out of the 75 days in the projection period fell above the 95% projection interval. In Scenarios A through D, the proportion increased from below 0.25 to close to 1 when increasing *σ* from 1 to 1.5, representing a 50% increase in the reinfection hazard coefficient.

**Figure 4.**
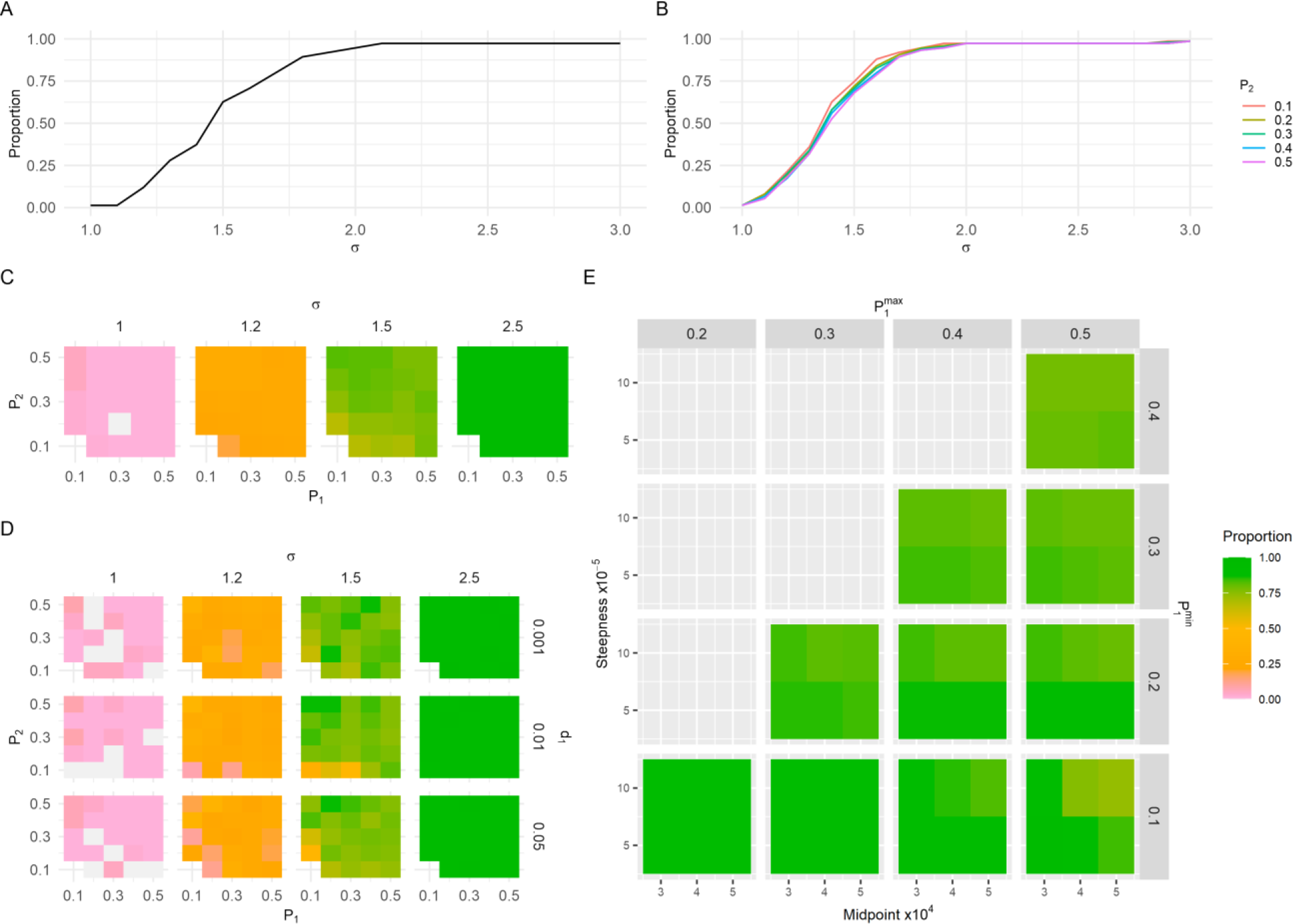
Median proportion of points above the projection interval after an increase in reinfection risk. Panels A-E represent Scenario A-E respectively, with scenario definitions as described in Figure 3. The gaps in C and D represent scenario definitions where results from all runs were excluded because of non-convergence or a cluster outside the projection interval during the fitting period.

In Scenarios A and B, the median proportion of points above the projection interval was 0.973 when *σ* ≥ 2.2 (Figure 4 A-B). There was a slight negative relationship between *P*_2_ and the proportion of points above the projection interval; the lower *P*_2_ values correlated with a higher proportion of points outside the projection interval) (Figure 4 B).

In Scenario C, when there was no change in reinfection risk (*σ* = 1) and the *P*_1_ values were low, there was a slightly higher proportion of points outside the projection interval compared to when *P*_1_ was higher (Figure 4 C). At *σ*_*t*_ = 1.5 (representing a 50% increase in the reinfection hazard coefficient), the proportion of points outside the projection interval was higher when more infections are observed. Even at a low observation probability, an increase in reinfection risk was still detected (*D*_*first*_ still existed). For instance, when *P*_1_ = 0.1 and *P*_2_ = 0.2, the median proportion of points above the projection interval was 0.67, whereas the proportion was closer to 0.8 in scenarios where *P*_1_ and *P*_2_ was higher. For substantial increases in the reinfection hazard coefficient (*σ* ≥ 1.7), the proportion of points above the projection interval was 0.973 (Figure S12).

The values used for mortality in Scenario D had a minimal effect on the proportion of points outside the projection interval (Figure 4 D), although the median proportion was slightly higher with higher *d*_1_ values.

In Scenario E, the median proportion of points above the projection interval varied from 0.01 to 0.25 when *σ* = 1 (indicating a 0% increase in the risk of reinfection), from 0.19 to 0.53 when *σ* = 1.2 and from 0.49 to 0.95 when *σ* = 1.5 (Figure 4 and Figure S13). The median proportion was lower at higher values of steepness for most scenario definitions, and at the higher value of midpoint values.

### Specificity

The specificity was 100% across all scenario definitions in Scenarios A and B. In Scenario C (Figure S14), where we considered fixed *P*_1_ and *P*_2_, and Scenario D (Figure S15), where we introduced mortality, the specificity was mostly above 0.95. However, in scenarios where *P*_1_ = 0.1 and the difference between *P*_1_ and *P*_2_ was substantial, such as *P*_1_ = 0.1 and *P*_2_ = 0.5, the specificity dropped to 0.75. In both Scenarios C and D, the specificity remained above 0.75.

In Scenario E, the specificity approached 1 when a larger number of cases were observed (i.e., higher values of 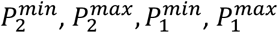, the midpoint, and steepness) (see Figure S17). Conversely, when fewer cases were observed, the specificity decreased. For instance, when 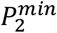 and 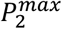 were 0.2 and 0.3 respectively, the specificity ranged from 0.33 to 0.91. Higher specificity values were observed when 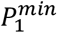 and 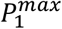 were higher, and the midpoint of the function was greater, indicating more cases were observed, as can been seen in Figure S17. When considering the runs where false positive increases in the reinfection risk were detected, most runs had a cluster of five consecutive observed reinfections below the interval during the fitting period, suggesting that the model did not align well with the trends in the data during the fitting period but did not match our exclusion criteria.

## Discussion

In this study, we performed simulation-based validation on a method used for real-time monitoring of SARS-CoV-2 reinfections to detect changes in the risk of reinfection (7). The model parameters converged well in most scenarios of observation process biases and the model is robust when dealing with changes in observation probability for reinfections. The robustness of the model’s parameter convergence indicates that the projection interval can be accurately simulated, particularly when the model aligns well with the patterns seen in the simulated data during the fitting procedure.

When both the observation probabilities for primary and reinfections were exceptionally low (0.1), convergence for the negative binomial dispersion parameter failed, due to a lack of data to properly inform this parameter. This finding aligns with a previous study by Pulliam *et al*. where the dispersion parameter did not converge over the short timeframe when fitting South African data over the first wave (18). Our findings further reinforce the importance of having sufficient data over a long-enough period to ensure accurate estimation of parameters.

In Scenario E, where we varied the observation probabilities based on the number of underlying primary infections using an adapted logistic function described, most runs converged successfully, except when the observed infections were low. In these cases, convergence of the dispersion parameter was sensitive to both the shape of the function and the number of runs that could converge. Convergence was lower when the observation probability decreased at a low number of underlying primary infections (midpoint of 30,000) and when the observation probability decreased gradually with increasing underlying primary infections (low steepness parameter). The non-convergence in scenario definitions with low observation probabilities is likely due to insufficient data to properly inform the negative binomial dispersion parameter.

We measured when the first cluster of five observed reinfections fell above the projection interval to understand how soon the approach detects changes in the reinfection risk and whether it detects changes when there were none (specificity). In all the data generation scenarios, there was a gradual decrease in the time it took to detect an increase in reinfection risk as the magnitude of the increase grew. In most scenarios, the increase in reinfection risk was detected even when the increase was as low as 20% and larger increases, above 50%, were detected soon after their introduction in the underlying data. However, when fewer infections and reinfections were observed, there was a slightly delayed detection in increase in reinfection risk which could be because of data availability during the fitting period, and even though the model parameters converged, the model parameters were not informed as well as when more cases were observed.

In evaluating the robustness of the approach’s ability to detect changes in the risk of reinfection, we introduced a parameter (σ ≥ 1) to scale the reinfection hazard coefficient used to generate reinfections after a certain date. The model-based detection approach was sensitive to changes in reinfection risk, detecting increases as low as 20% (σ ≥ 1.2) when considering imperfect observation of both primary infections and reinfections. The detection of an increase in reinfection risk at a low magnitude highlights the method’s sensitivity.

Furthermore, when the observation probabilities were varied as a function of underlying primary infections, the proportion of observed reinfections above the projection interval for a given magnitude of increase in the reinfection hazard coefficient remained consistent despite changes in the function’s parameters. For instance, when reinfection risk increased by 50%, more than half (above 0.5) the observed reinfections fell above the projection interval, indicating that the method is sensitive to increases in the reinfection risk.

We also evaluated specificity, measuring the proportion of scenarios where no increase in reinfection risk was detected (i.e., there were no stretches of five consecutive points above the projection interval), given that no such increase was present in the simulated data (where σ = 1). In scenarios where all primary infections were observed (Scenarios A and B), there were no false positive detections of changes in reinfection risk. However, when observation probabilities for primary infections were included alongside reinfections, there were some runs where false increases in the risk of reinfections were detected, particularly when the difference between observation probabilities for primary infections and reinfections was high (>0.3).

When observation probabilities were calculated as a function of the number of underlying primary infections, few false positives were detected when the number of observed cases were high. However, when the infections and reinfections observed are lower, especially when the observed infections did not fall well within the projection interval during the fitting period (i.e., there are large clusters below or above the projection interval), we advise more careful interpretation of apparent changes in the risk of reinfection. In such cases, criteria to detect an increase in the risk of reinfection could, for example, be extended to having a cluster of ten consecutive days above the projection interval during the projection period instead of five.

Notably, changes in mortality had negligible effects on all performance metrics, including convergence, the proportion of points above the projection interval, the first cluster of five observed reinfections above the projection interval and specificity. This indicates the method’s reliability even when mortality rates fluctuate.

Overall, the simulation-based validation of this catalytic model for detecting SARS-CoV-2 reinfection risk provides useful insights on how the model should be used as a tool when monitoring reinfection risk to respond to increases in the reinfection risk.

## Strengths and limitations

A major strength of this study is that we investigated the robustness of the model under different assumptions of observation probabilities that could occur in the real-world, enhancing the model’s practical applicability. Such robustness ensures that the model remains reliable when introduced to various complex scenarios in the real-world.

Recognising that mortality may bias the number of people eligible for reinfection, we incorporated mortality and determined that the model outcome is not sensitive to changes in mortality rates which could be influenced in the real world by factors such as healthcare capacity, treatment effectiveness, and vaccination campaigns.

A key limitation of this study is the timeframe on which the work has been conducted. Ideally, this type of simulation-based validation should be conducted before such a method is used for real-time monitoring during an outbreak response. While the method is intended for real-time detection in changes in the risk of reinfection by SARS-CoV-2; resources were not available to conduct the simulation-based validation to coincide with the SARS-CoV-2 outbreak response.

Another limitation is that the simulated dataset used in the simulation-based validation is based on the situation in South Africa; thus, the findings may not be applicable to countries with significantly small populations, limited testing, or extensive vaccination coverage, resulting in low numbers of observed infections and reinfections.

Lastly, the simulation-based validation did not consider waning natural immunity as a potential reason for an increase in the risk of reinfection. The method focuses on detecting a population-level increase in the reinfection risk but does not assign a mechanism to the detected increase; interpretation of the drivers of a change in reinfection risk requires triangulation with other data sources. That said, whilst there is evidence of waning natural immunity of SARS-CoV-2 (19), analysis of reinfection trends in South Africa was conducted from January 2021 through November 2022, with the only detected change in reinfection risk being associated with the emergence of the Omicron variant. This finding suggests that the dynamics of waning immunity for SARS-CoV-2 may not produce population-level increases in reinfection risk that are detectable using this method.

## Directions for future work

The data used to validate the model in this paper (7), is based on a population that represents South Africa. Since we have found that with a low number of cases, the model parameters may fail to converge, research is also needed to validate the model with data representative of different countries, population sizes, and vaccination histories to ensure its broader applicability.

Vaccination against SARS-CoV-2 is increasing globally and may have an impact in reinfection risk. Future work should therefore study the impact of vaccination on the robustness of the model and may require modification of the method for use in contexts where vaccination levels are high and/or changing at a fast pace.

Pulliam *et al*. used a second approach to detect changes in the risk of reinfection, which estimated time-varying infection and reinfection hazards (7). Like this study, simulation-based validation should be performed for this method.

Since we did not consider waning immunity in the simulation-based validation, it remains to be seen whether and under what conditions waning immunity would produce a population-level shift in reinfection risk that would be detected by this method.

## Conclusions

Simulation-based validation has been conducted on the method that uses a catalytic model to detect increases in the risk of reinfection by SARS-CoV-2. Although continued validation under different epidemiological contexts is necessary, the work done in this study demonstrates the method’s robust performance across most imperfect observation and mortality scenarios. Specifically, model parameter convergence and good fit during the fitting period should be prerequisites when using the model to detect real-time increases in population-level reinfection risk. The simulation-based validation done on the catalytic model enhances the model’s applicability when using the model to draw conclusions under different scenarios that might occur in real-world data.

## Data Availability

All data produced are available online at https://github.com/SACEMA/reinfectionsBelinda and https://zenodo.org/record/8354838 (supplemented by https://zenodo.org/record/7426515)

## Acknowledgements

J.R.C.P. and C.v.S. are supported by the South African Department of Science and Innovation and the National Research Foundation. Any opinion, finding, and conclusion or recommendation expressed in this material is that of the authors, and the NRF does not accept any liability in this regard. This work was also supported by the Wellcome Trust (grant no. 221003/Z/20/Z) in collaboration with the Foreign, Commonwealth and Development Office, United Kingdom. Writing assistance was provided by Yuri Munsamy, PhD of SACEMA, South Africa. This assistance was funded by Stellenbosch University. The authors gratefully acknowledge the Centre for High Performance Computing (CHPC), South Africa, for providing computational resources to this research project.

This work has benefited from input during the Clinic on Meaningful Modeling of Epidemiological Data (MMED) and the Software for the Applied Mathematical Sciences (SEAMS) workshop, both of which are part of the International Clinics on Infectious Disease Dynamics and Data (ICI3D) program. We specifically thank C. A. B. Pearson, T.J. Hladish, A. Stoltzfus, S. Horn, Y. Jo, L. S. Villabona-Arenas for helpful discussions during the development of this work.

## Supplementary Material

### Simulated Data

**Figure S1.**
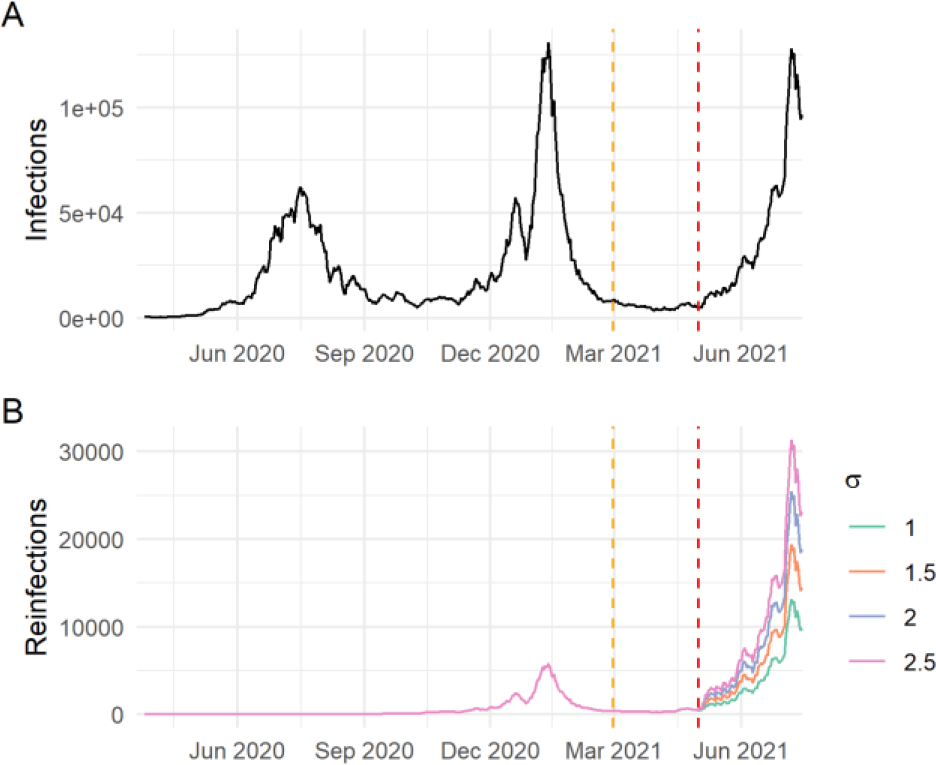
Simulated underlying primary infections with reinfections at different values of σ. The plot represents Scenario A, with figure A showing the simulated primary infections with perfect observation and no mortality, and B showing the observed reinfections with different values of σ used as input in Scenario A.

**Figure S2.**
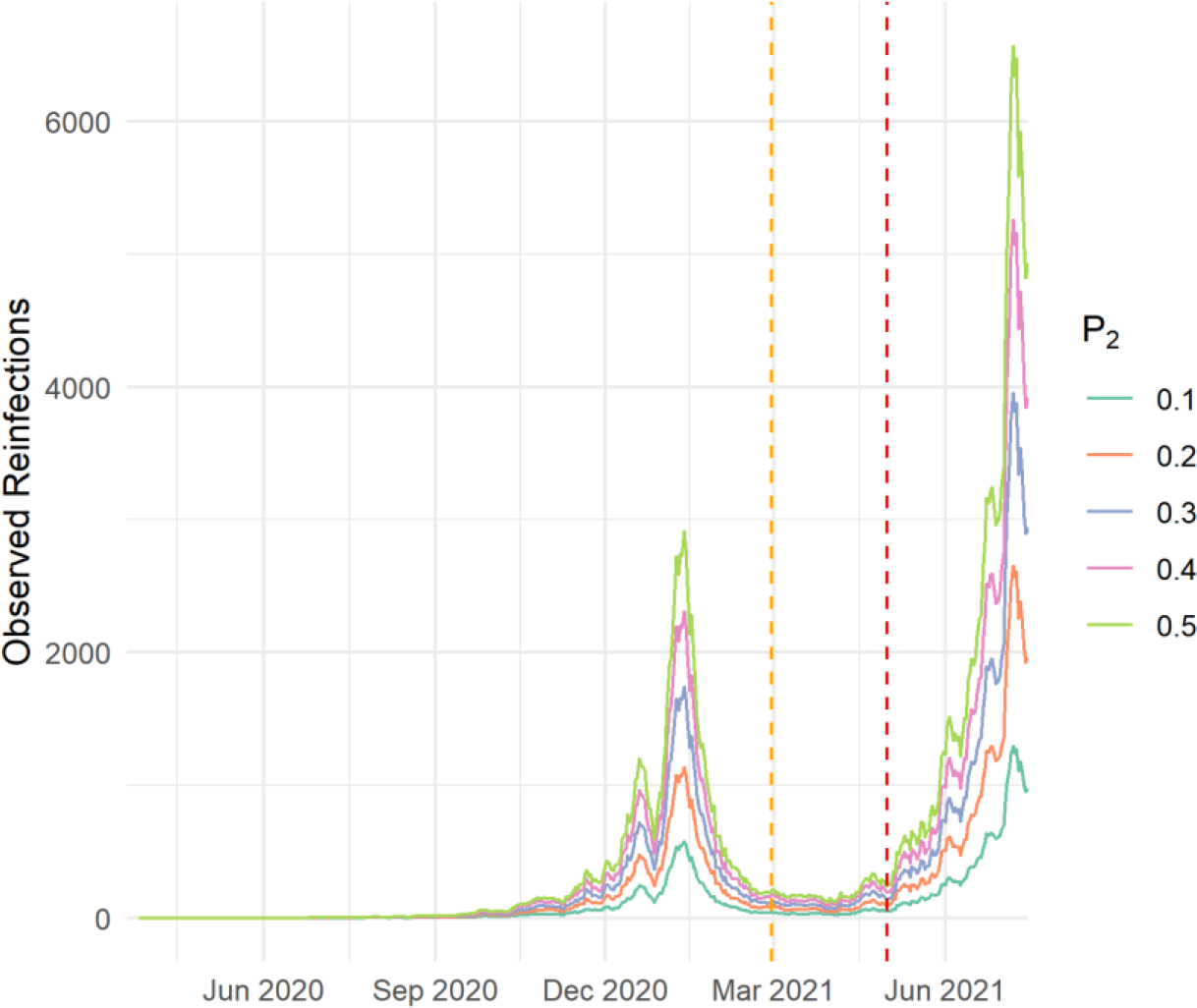
Observed reinfections for different observation probabilities for reinfections, P_2_, in Scenario B (no change in reinfection risk) and σ = 1. Scenario B has imperfect observation of reinfections.

**Figure S3.**
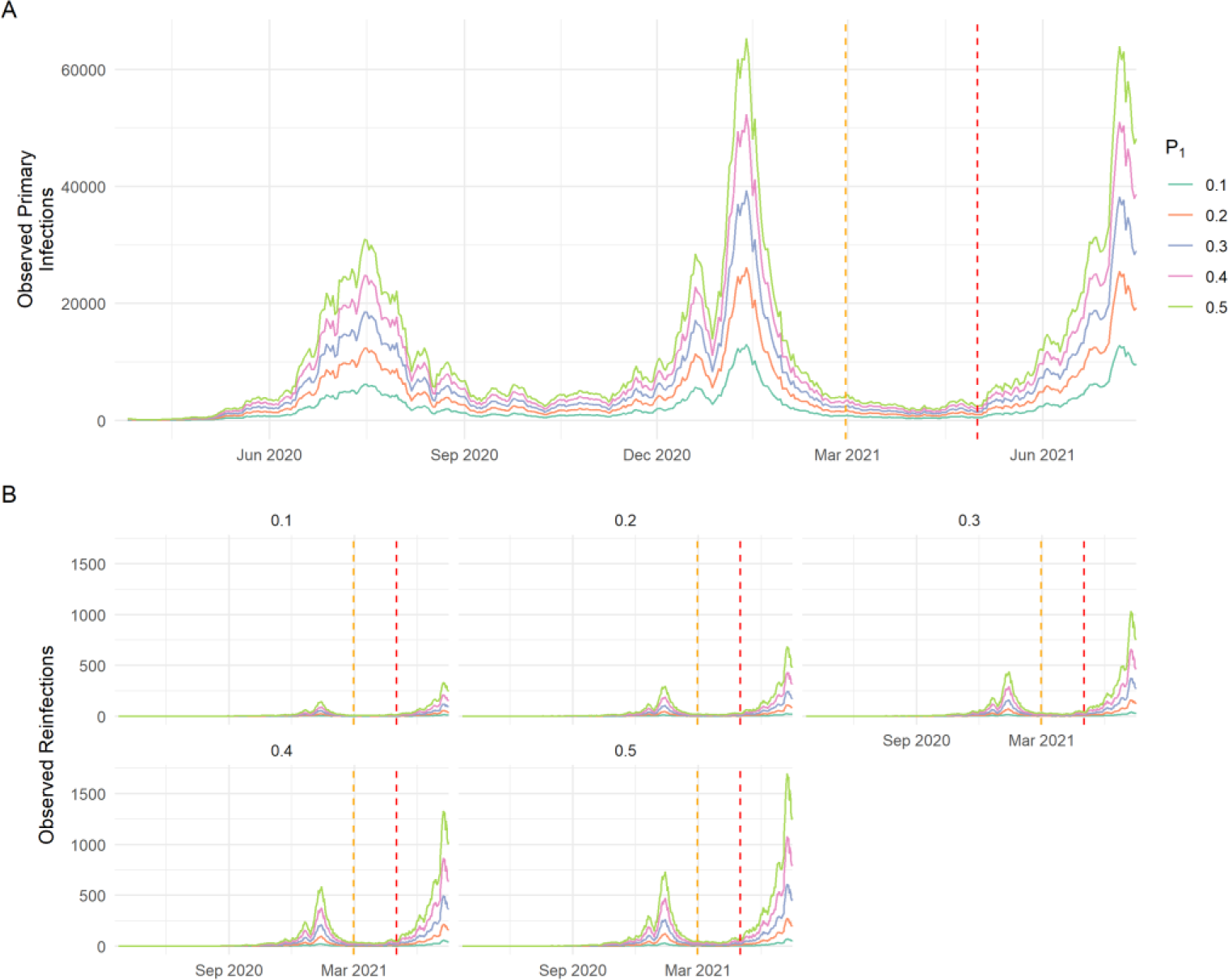
The observed primary infections and reinfections for Scenario C. A shows the number of observed primary infections for different values of P_1_and B shows the observed reinfections for different values of P_2_shown at the top of each grid. Each line depicts another value of P_1_.

### Parameter convergence

**Figure S4.**
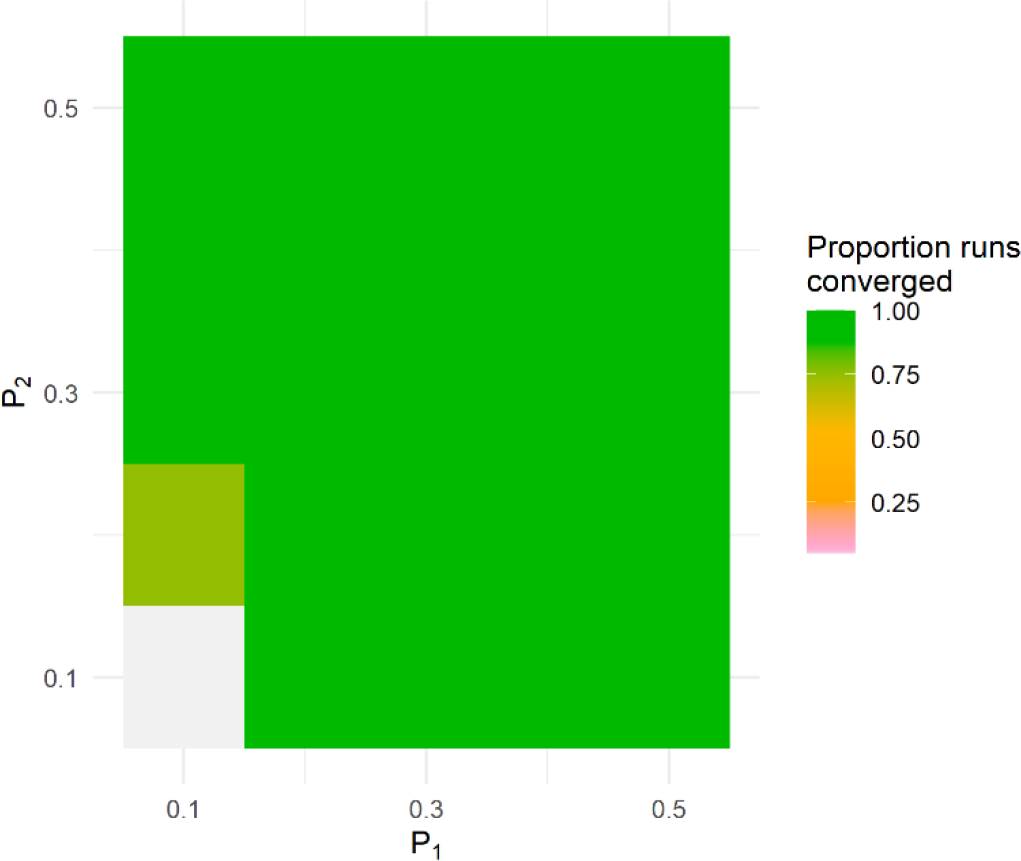
Proportion of runs in Scenario C where both λ and κ converged. Here we introduced observation probabilities for primary infections and reinfections (P_1_and P_2_respectively).

**Figure S5.**
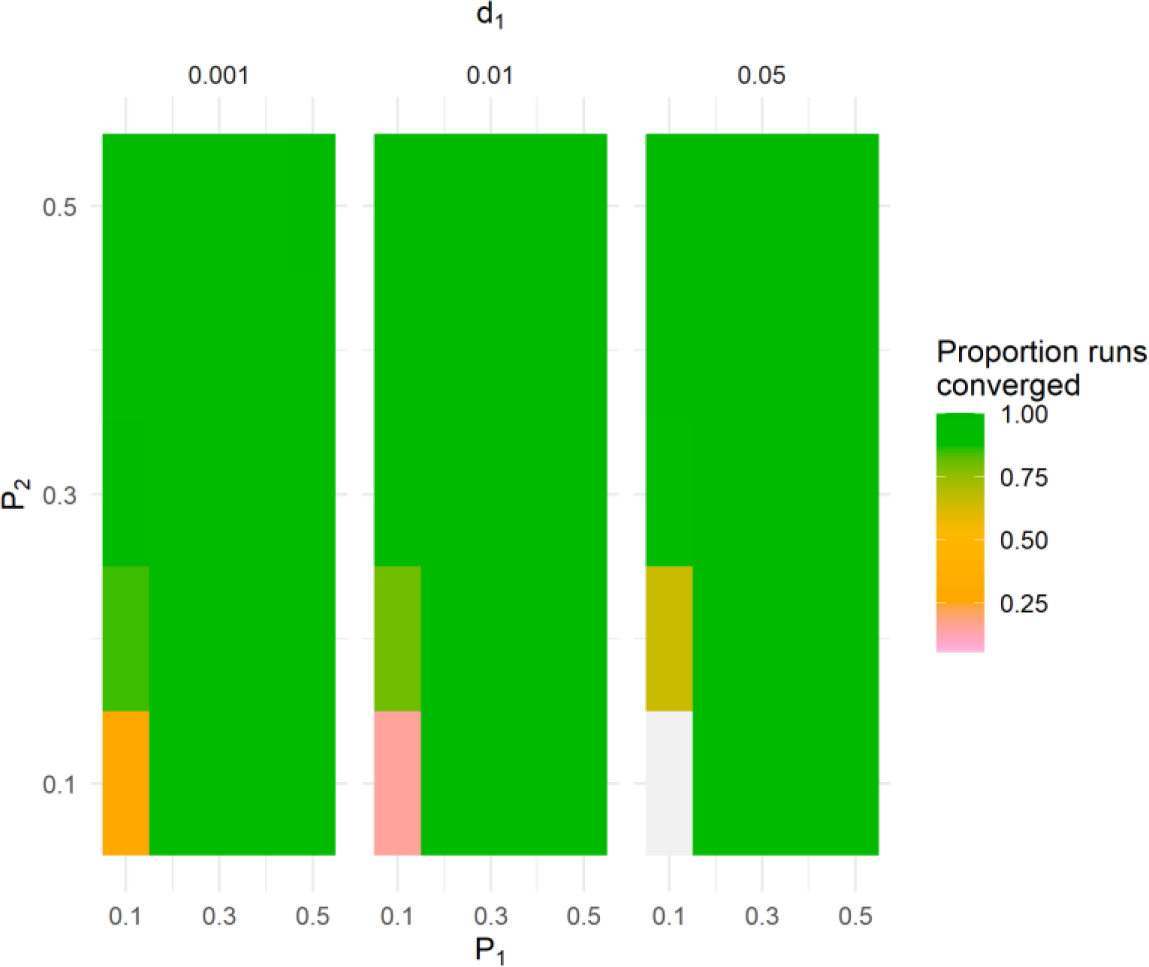
Proportion of runs in Scenario D where both λ and κ converged. Here we added observation probabilities for primary infections, reinfections and we included mortality (P_1_, P_2_and d_1_respectively).

**Figure S6.**
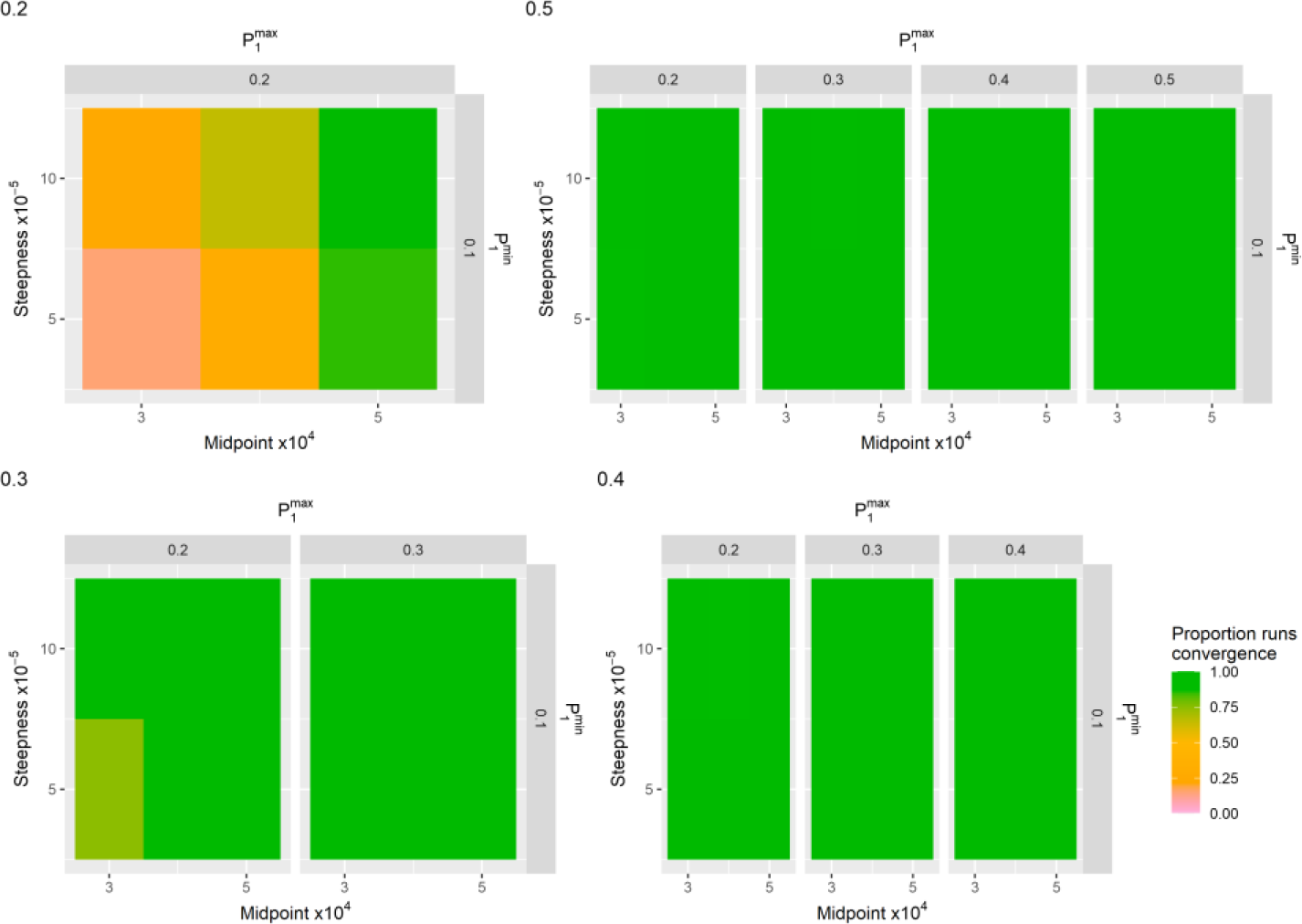
The proportion of runs that converged for Scenario E where 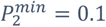 and 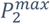 indicated at the top of each grid.

**Figure S7.**
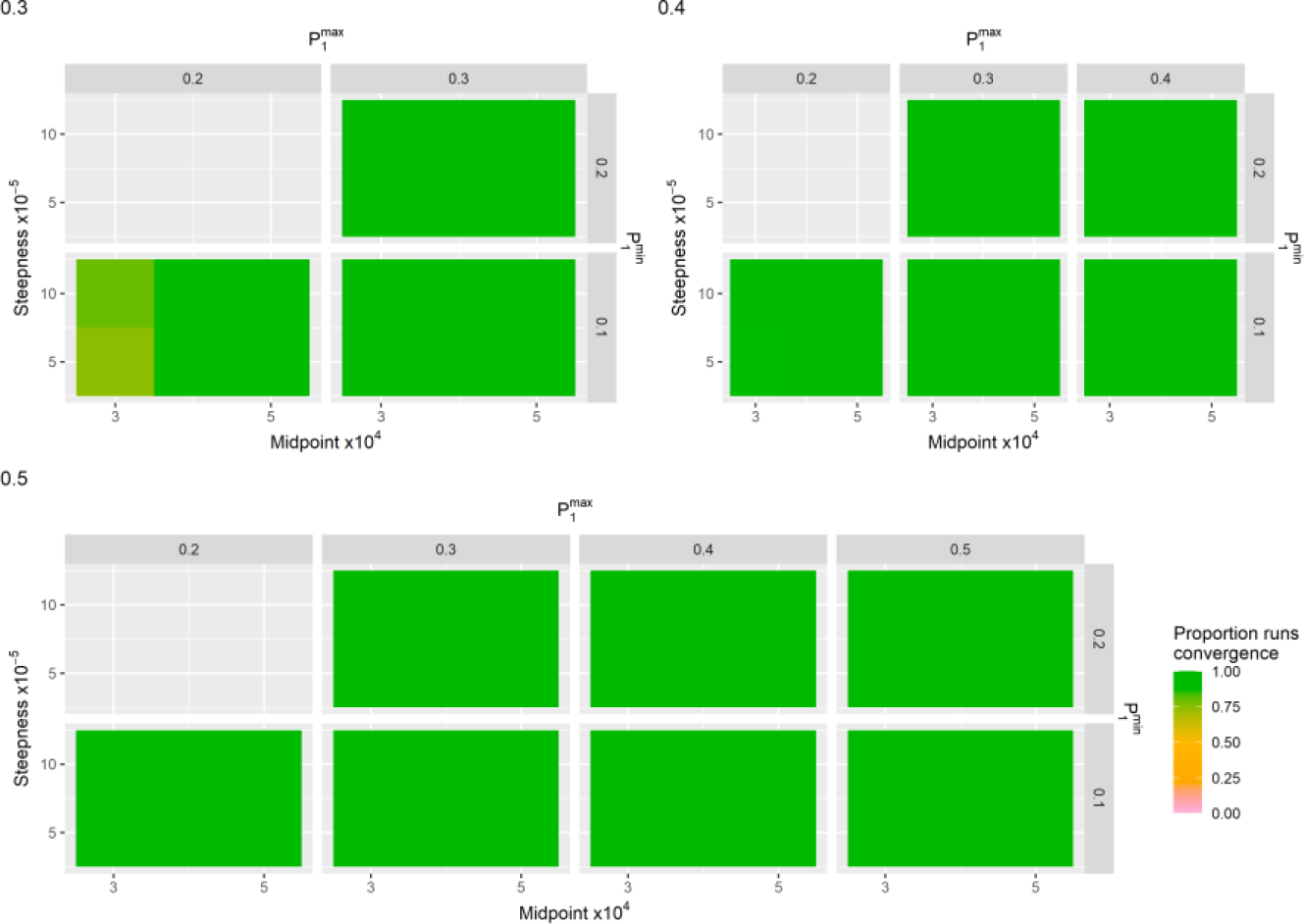
The proportion of runs that converged for Scenario E where 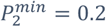 and 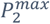 indicated at the top of each grid.

**Figure S8.**
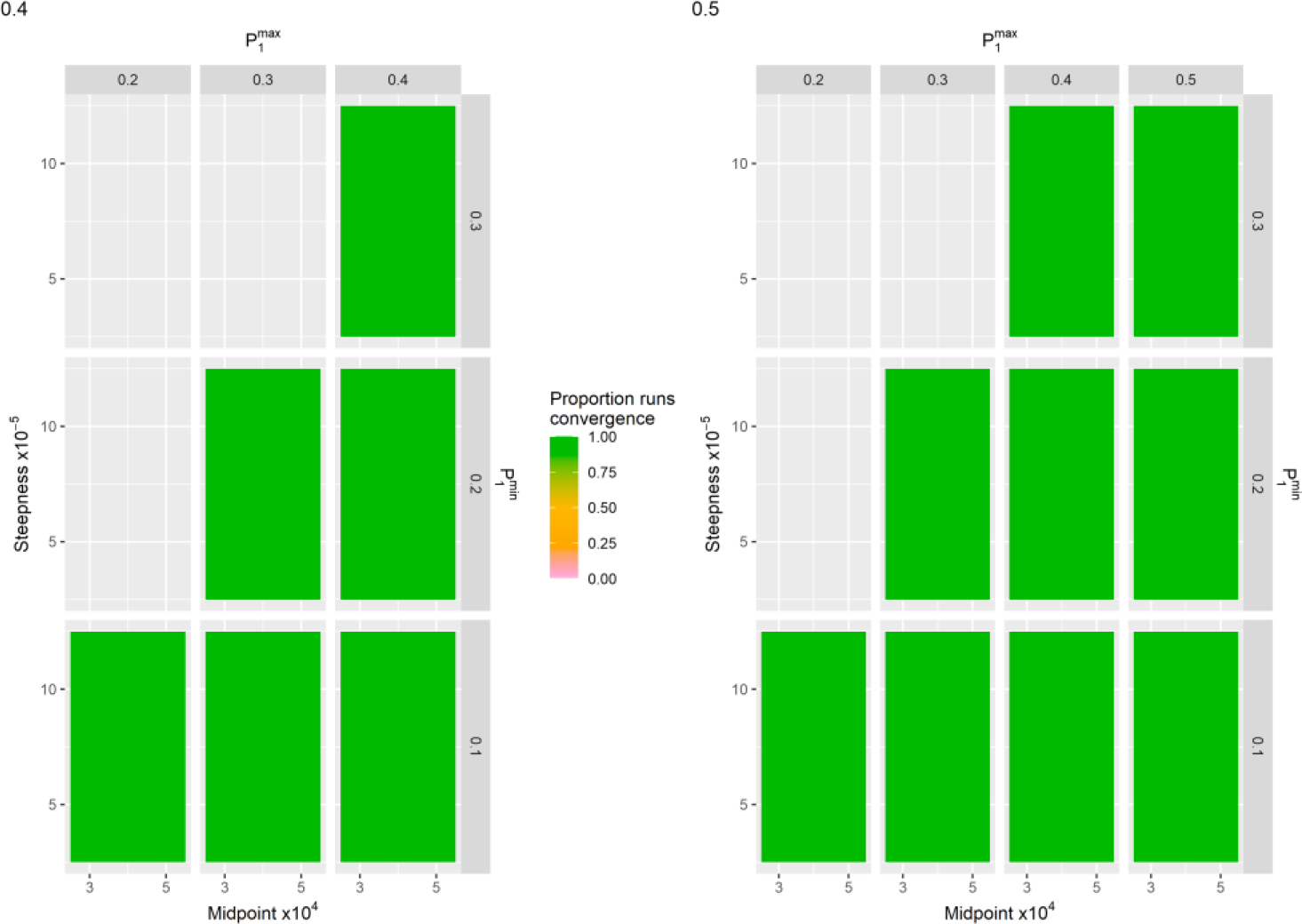
The proportion of runs that converged for Scenario E where 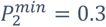 and 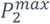 indicated at the top of each grid.

**Figure S9.**
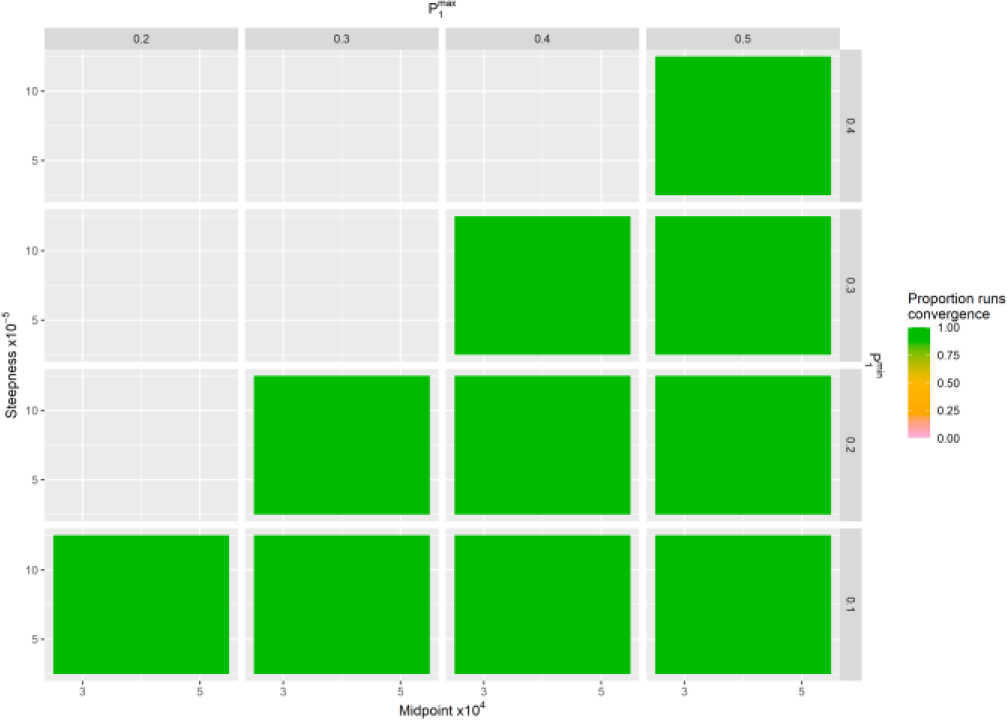
The proportion of runs that converged for Scenario E where 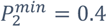 and 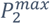 indicated at the top of each grid.

### Exclusion of non-converging runs or poor model fit

**Figure S10.**
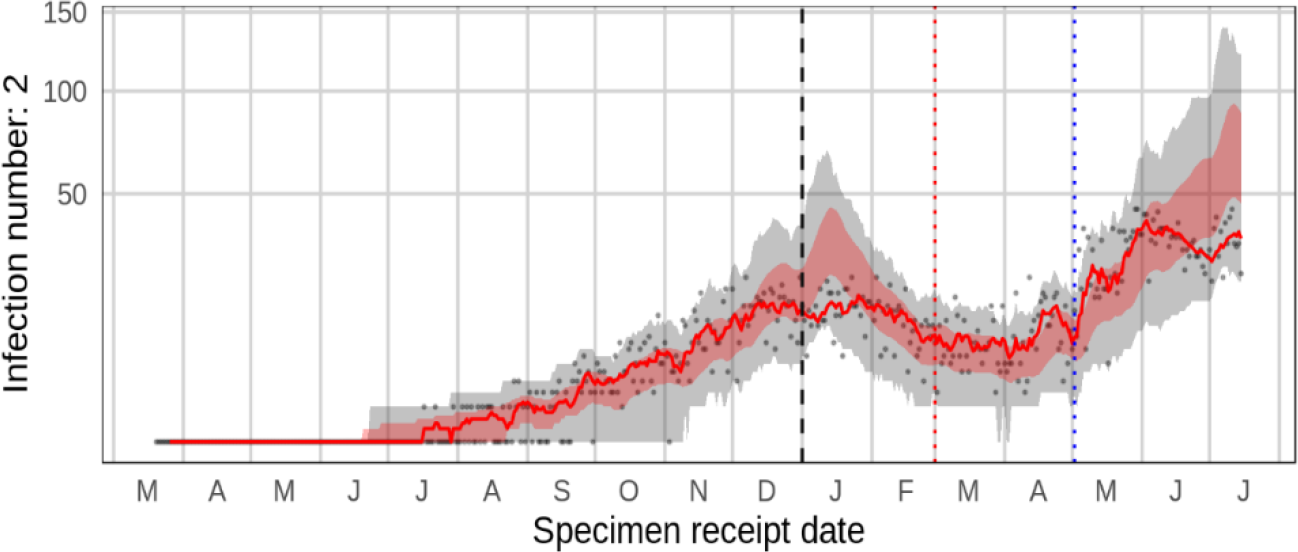
An instance of an ‘unrealistic’ time series where the model faced challenges in fitting the simulated reinfection data. During the fitting period preceding the dotted red line, the observed reinfections (depicted by the solid red line) consistently fell below the projection interval in January. In this particular scenario, the function determining the observation probability had a low midpoint of 30,000, minimal observation probabilities for primary and re-infections set at 0.1 and 0.4, respectively, and a low steepness factor of 0.00005.

### First cluster of reinfections above the projection interval

**Figure S11.**
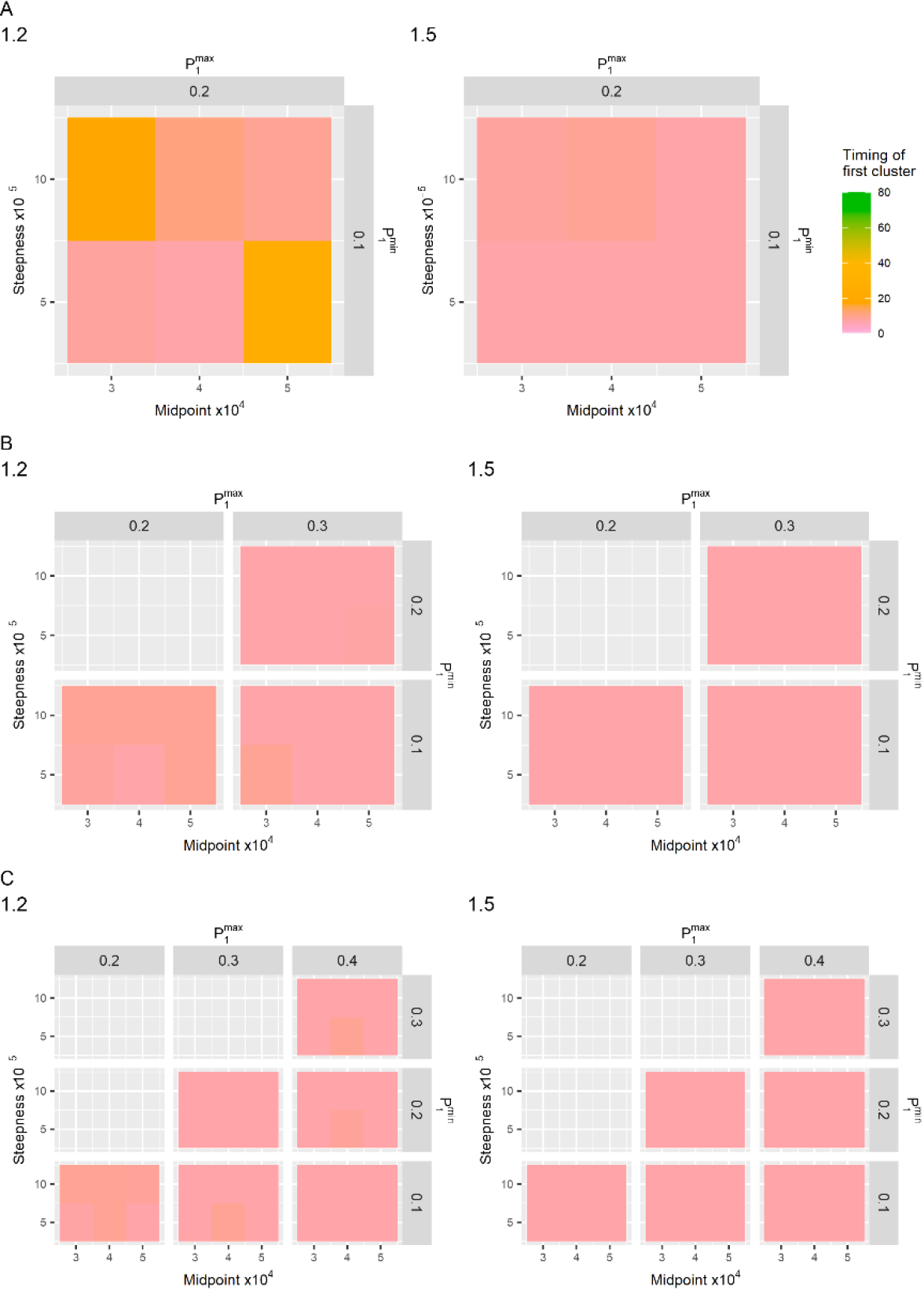
Plot showing the median of the timing of the first cluster of five days where the reinfections fell above the projection interval after the introduction of the scale (σ) for Scenario E. In A, the minimum and maximum observation probabilities for reinfections are 0.1 and 0.2. In B, the minimum and maximum observation probabilities for reinfections are 0.2 and 0.3. In C, the minimum and maximum observation probabilities are 0.3 and 0.4. The introduced scales (σ) are indicated at the top.

### Proportion of infections above projection interval

**Figure S12.**
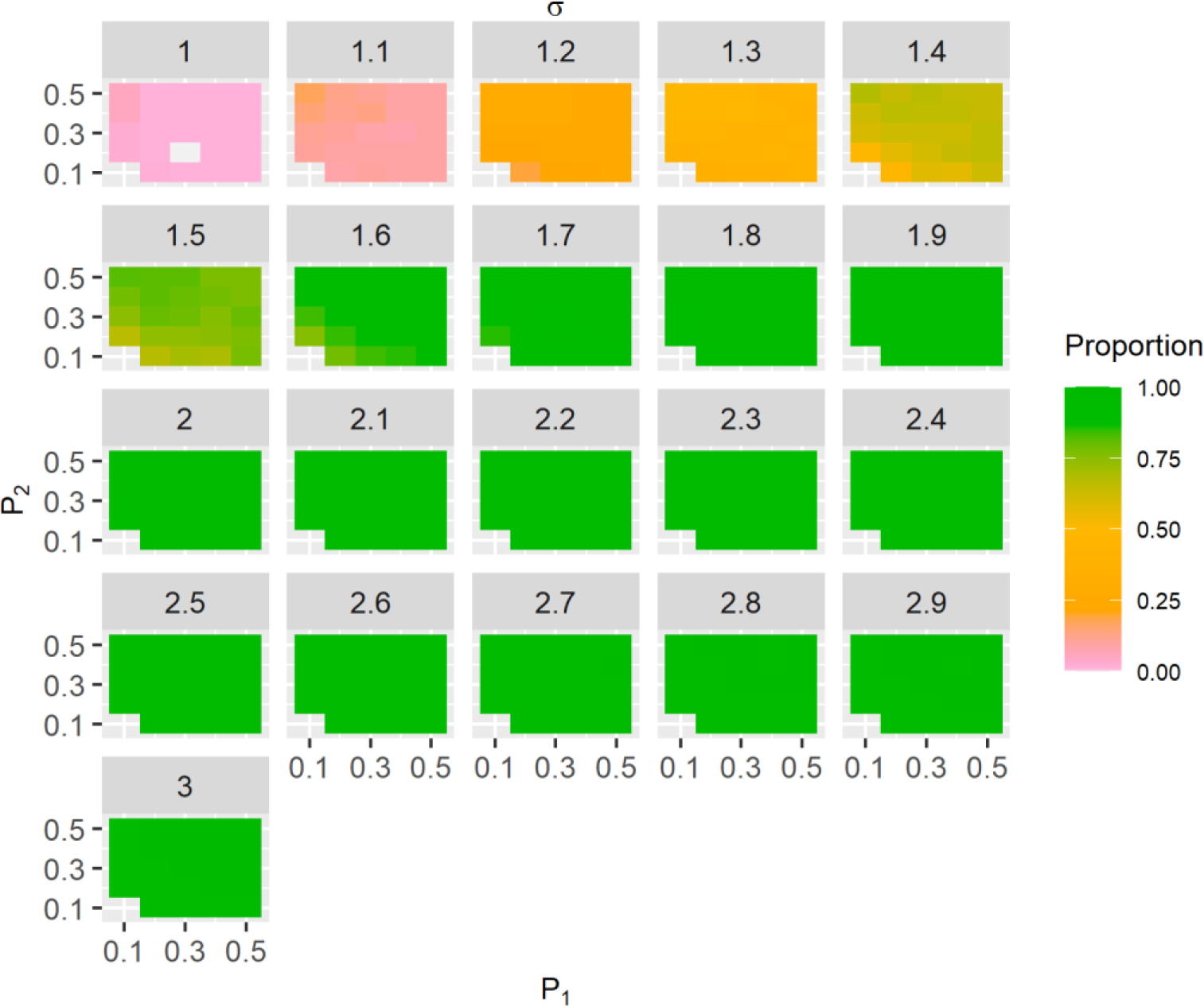
Plot showing the median of the proportion of points above the projection interval for Scenario C for different values σ.

**Figure S13.**
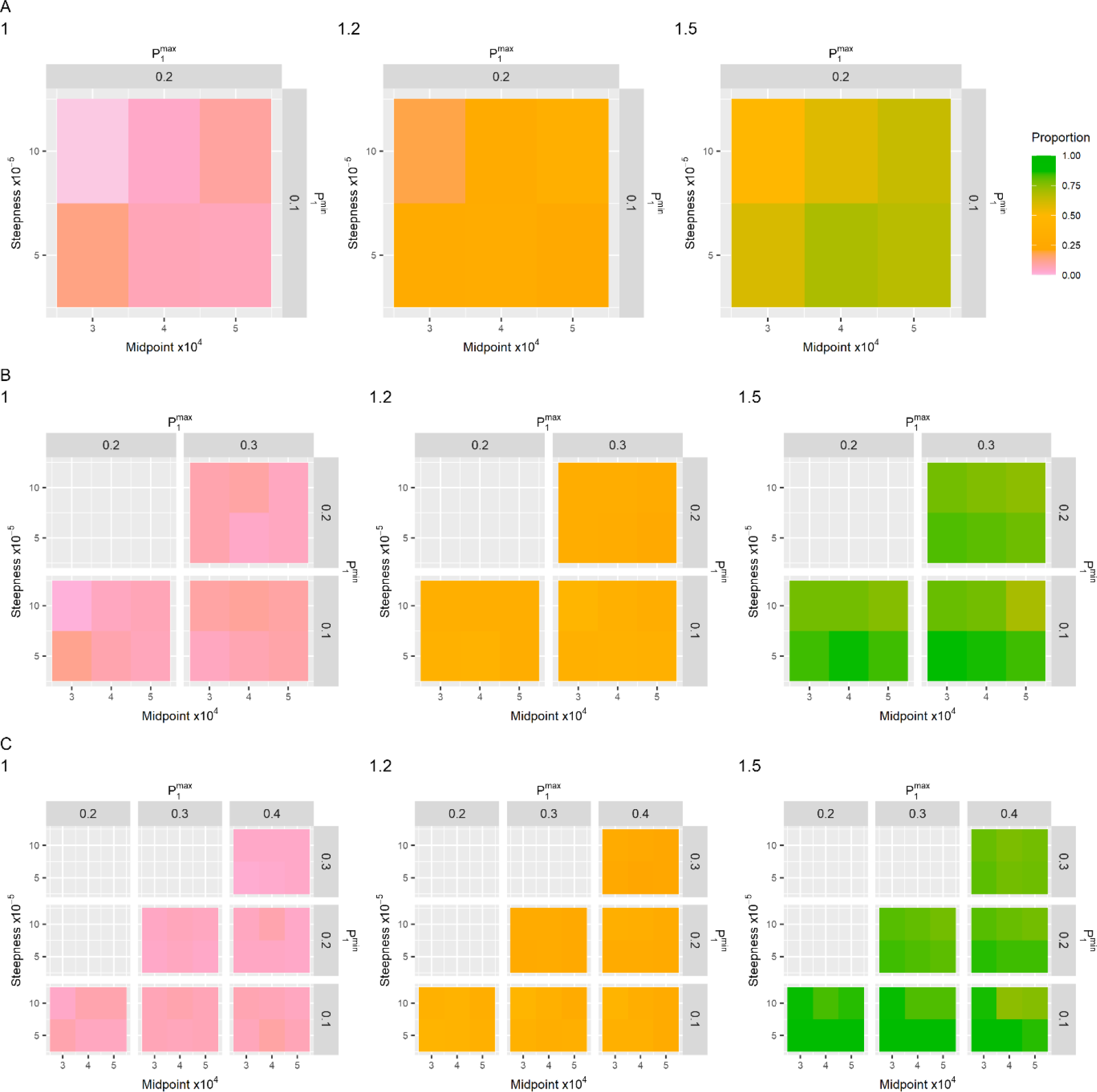
Plot showing the median of the proportion of points above the projection interval for Scenario E. In A, the minimum and maximum observation probabilities for reinfections are 0.1 and 0.2. In B, the minimum and maximum observation probabilities for reinfections are 0.2 and 0.3. In C, the minimum and maximum observation probabilities are 0.3 and 0.4. The introduced scales (σ) are indicated at the top.

### Specificity

**Figure S14.**
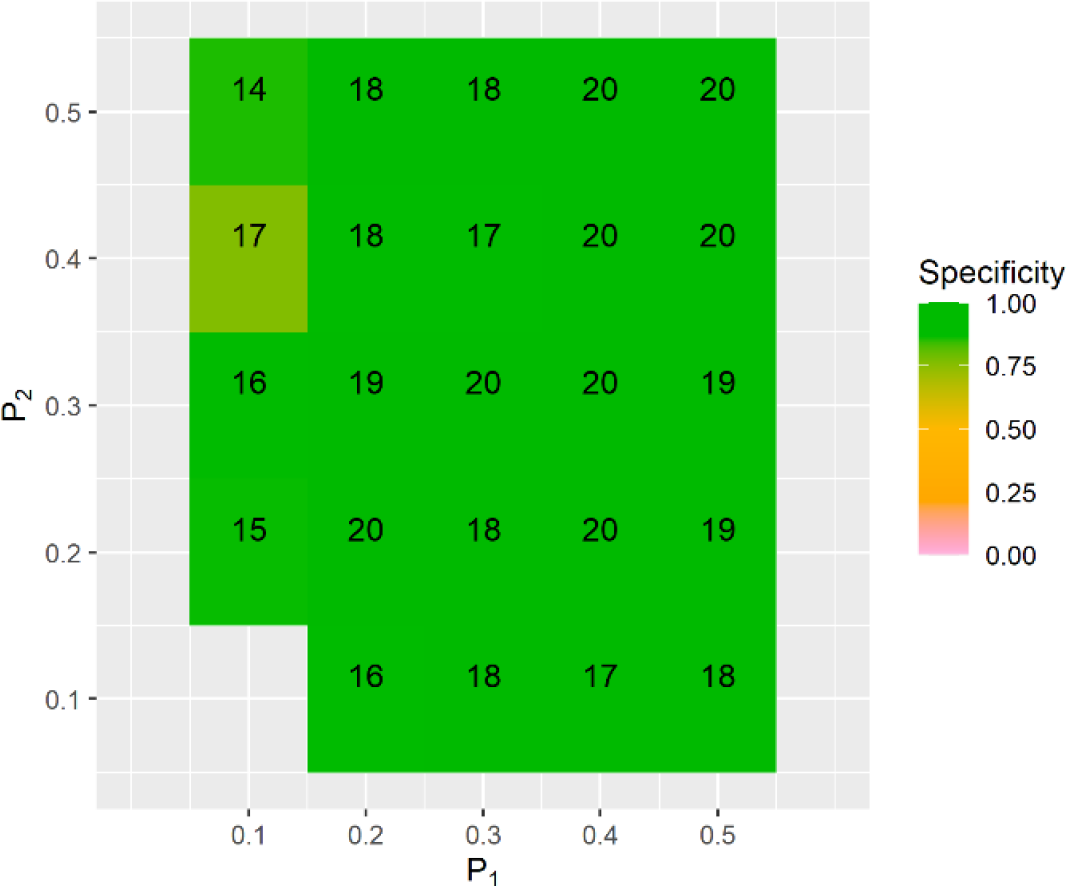
Specificity (σ = 1) of Scenario C, over 20 runs with different fixed values of primary infections and reinfections observation probabilities. The numbers in the grid are the number of runs where both λ and κ converged and a cluster of five consecutive points above or 10 consecutive points below the projection interval during the fitting period does not exist. The specificity is measured as the number of those runs where D_first_ does not exist, i.e., no false positive detection of a change in reinfection risk was observed.

**Figure S15.**
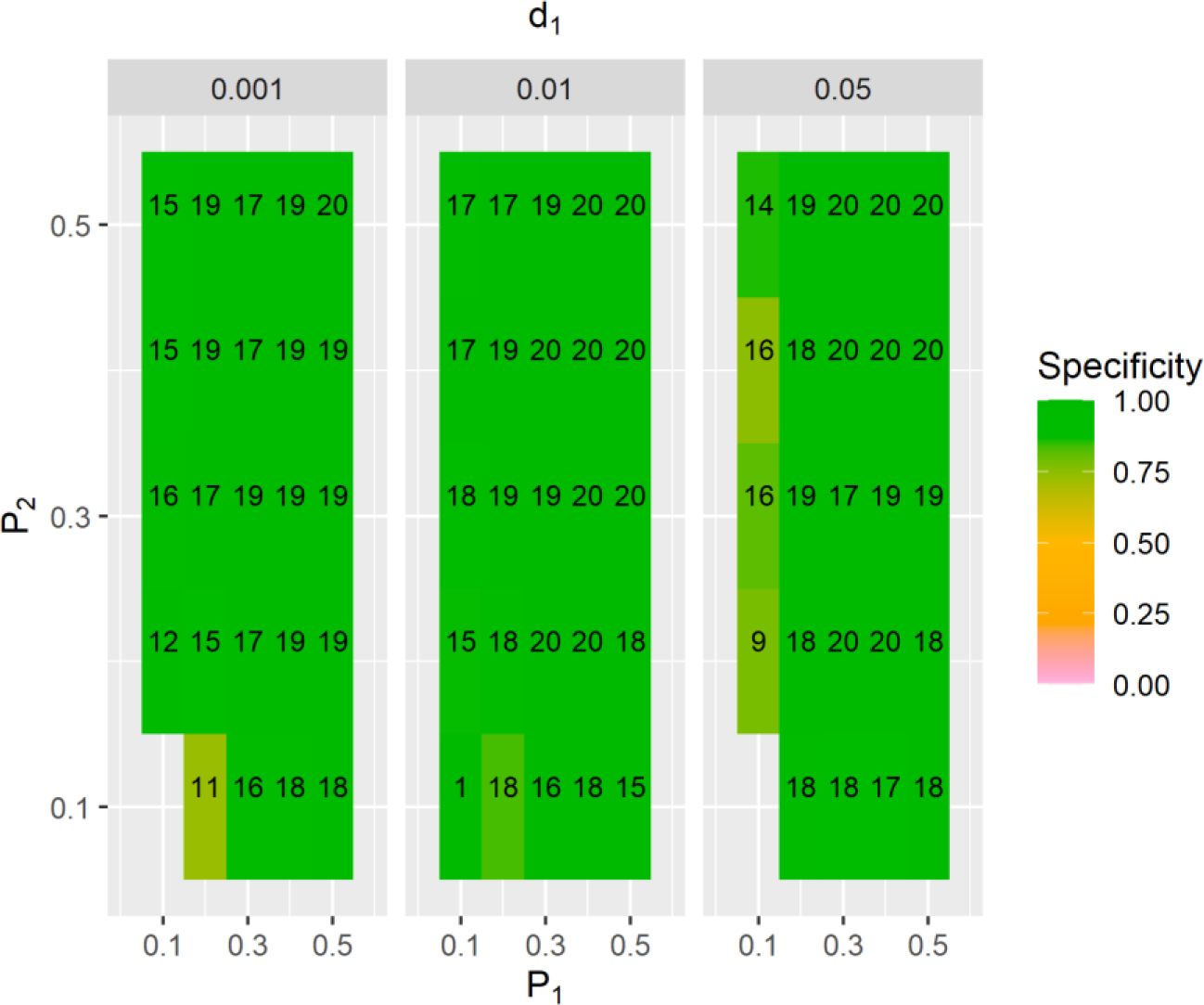
Specificity (σ = 1) of Scenario D over 20 runs with different fixed values of primary infections and reinfections observation probabilities where mortality is considered. The numbers in the grid are the number of runs where both λ and κ converged and a cluster of five consecutive points above or 10 consecutive points below the projection interval during the fitting period does not exist. The specificity is measured as the number of those runs where D_first_ does not exist, i.e., no false positive detection of a change in reinfection risk was observed.

**Figure S16.**
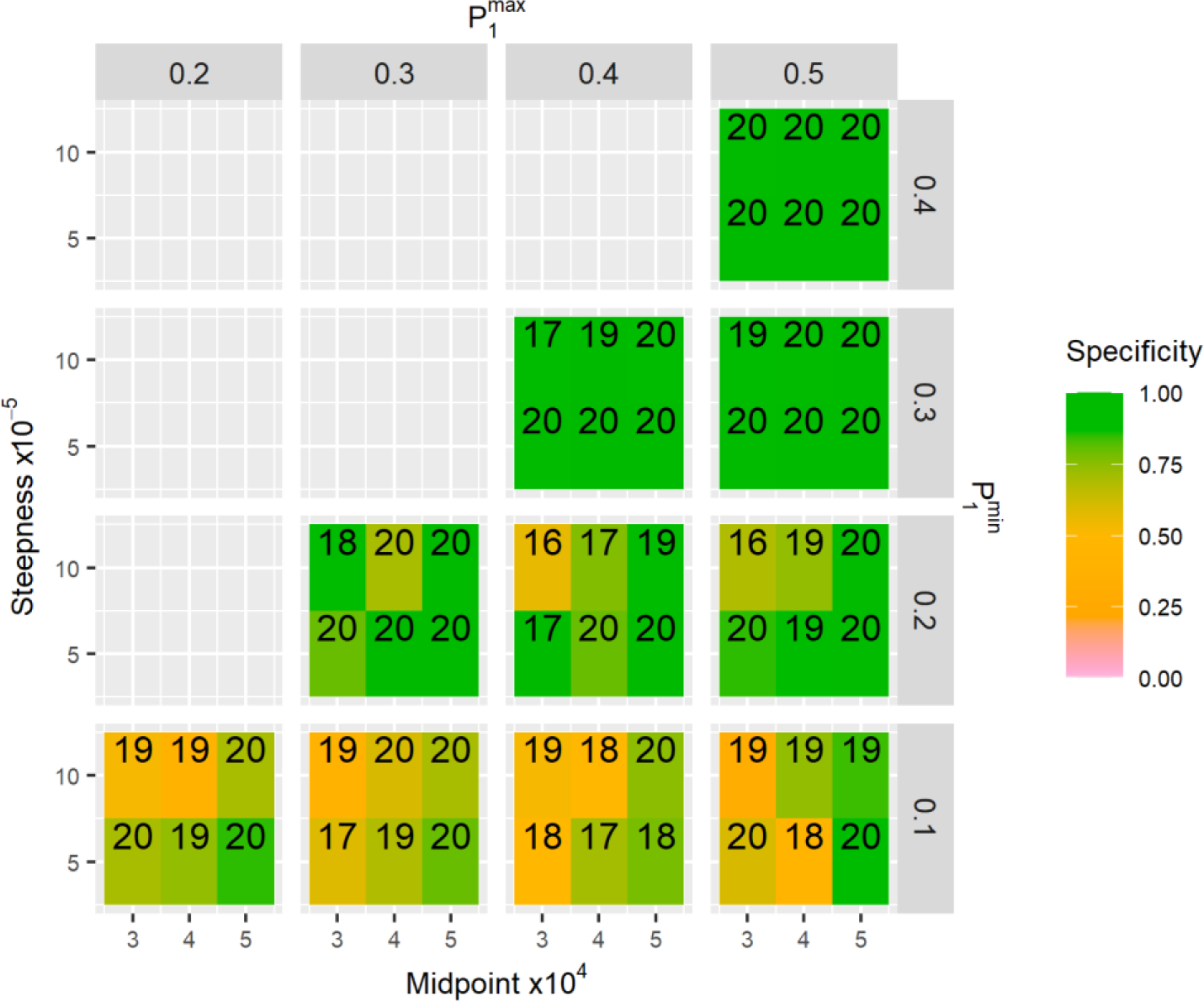
Specificity (σ = 1) of Scenario E over 20 runs when 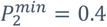 and 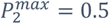. The numbers in the grid are the number of runs where both λ and κ converged and a cluster of five consecutive points above or 10 consecutive points below the projection interval during the fitting period does not exist. The specificity is measured as the number of those runs where D_first_ does not exist, i.e., no false positive detection of a change in reinfection risk was observed.

**Figure S17.**
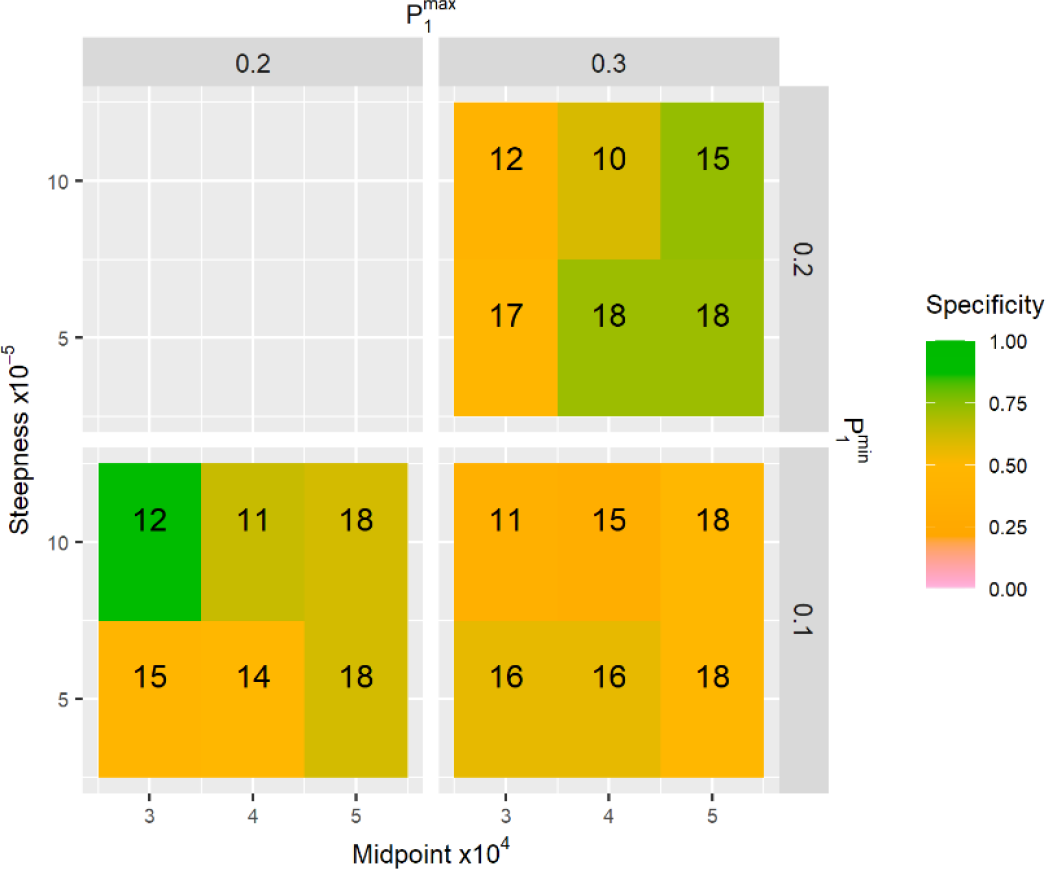
Specificity (σ = 1) of Scenario E over 20 runs when 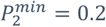 and 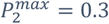. The numbers in the grid are the number of runs where both λ and κ converged and a cluster of five consecutive points above or 10 consecutive points below the projection interval during the fitting period does not exist. The specificity is measured as the number of those runs where D_first_ does not exist, i.e., no false positive detection of a change in reinfection risk was observed.

## References

1. WHO Coronavirus (COVID-19) Dashboard [Internet]. [cited 2023 Jul 18]. Available from: https://covid19.who.int/

2. Tegally H, Moir M, Everatt J, Giovanetti M, Scheepers C, Wilkinson E, et al. Emergence of SARS-CoV-2 Omicron lineages BA.4 and BA.5 in South Africa. Nat Med. 2022;28(28).

3. Bingham J, Cable R, Coleman C, Glatt TN, Grebe E, Mhlanga L, et al. Estimates of prevalence of anti-SARS-CoV-2 antibodies among blood donors in South Africa in March 2022. Res Sq. 2022;

4. Hernandez-Suarez C, Murillo-Zamora E. Waning immunity to SARS-CoV-2 following vaccination or infection. Front Med (Lausanne). 2022;9.

5. Atifa A, Khan MA, Iskakova K, Al-Duais FS, Ahmad I. Mathematical modeling and analysis of the SARS-Cov-2 disease with reinfection. Comput Biol Chem. 2022;98.

6. Coutinho RM, Marquitti FMD, Ferreira LS, Borges ME, da Silva RLP, Canton O, et al. Modelbased estimation of transmissibility and reinfection of SARS-CoV-2 P.1 variant. Communications Medicine. 2021;1(1).

7. Pulliam JRC, van Schalkwyk C, Govender N, von Gottberg A, Cohen C, Groome MJ, et al. Increased risk of SARS-CoV-2 reinfection associated with emergence of Omicron in South Africa. Science (1979). 2022;376(376).

8. Gibbons CL, Mangen MJJ, Plass D, Havelaar AH, Brooke RJ, Kramarz P, et al. Measuring underreporting and under-ascertainment in infectious disease datasets: A comparison of methods. BMC Public Health. 2014;14(14).

9. Lau H, Khosrawipour T, Kocbach P, Ichii H, Bania J, Khosrawipour V. Evaluating the massive underreporting and undertesting of COVID-19 cases in multiple global epicenters. Pulmonology. 2021;27(27).

10. Rahmandad H, Lim TY, Sterman J. Behavioral dynamics of COVID-19: estimating underreporting, multiple waves, and adherence fatigue across 92 nations. Syst Dyn Rev. 2021;37(37).

11. Banerjee I, Robinson J, Sathian B. COVID-19: Are reinfections a global health threat? Nepal J Epidemiol. 2021;11(11).

12. Robinson S. Simulation verification, validation and confidence: A tutorial. Transactions of the Society for Computer Simulation. 1999;16(16).

13. Wei Y, Sha F, Zhao Y, Jiang Q, Hao Y, Chen F. Better modelling of infectious diseases: Lessons from covid-19 in China. The BMJ. 2021;375.

14. Sargent RG. Verification and Validation of Simulation Models: An Advanced Tutorial. In: Proceedings - Winter Simulation Conference. 2020.

15. Pulliam JRC, van Schalkwyk C, Govender N, von Gottberg A, Cohen C, Groome MJ, et al. Data for Increased risk of SARS-CoV-2 reinfection associated with emergence of Omicron in South Africa. 2022 Feb 16 [cited 2023 Jul 20]; Available from: https://zenodo.org/record/6108448

16. Team RC. R: A Language and Environment for Statistical Computing. R Foundation for Statistical Computing.2021.

17. Gelman A, Rubin DB. Inference from iterative simulation using multiple sequences. Statistical Science. 1992;7(7).

18. Brooks SP, Gelman A. General methods for monitoring convergence of iterative simulations)? Journal of Computational and Graphical Statistics. 1998;7(7).

19. Xiang T, Liang B, Fang Y, Lu S, Li S, Wang H, et al. Declining Levels of Neutralizing Antibodies Against SARS-CoV-2 in Convalescent COVID-19 Patients One Year Post Symptom Onset. Front Immunol. 2021 Jun 16;12:708523.

